# Network topology outweighs emergence probability in surveillance sentinel placement

**DOI:** 10.1101/2025.11.10.25339713

**Authors:** Xiaolu Wang, Victor Del Rio Vilas, Qi Li, Qu Cheng

**Affiliations:** Department of Epidemiology and Biostatistics, School of Public Health, Tongji Medical College, Huazhong University of Science and Technology, Wuhan, Hubei, People’s Republic of China; UK-Public Health Rapid Support Team, UK Health Security Agency, London, United Kingdom

## Abstract

**Background:** Many studies have focused on understanding spatial heterogeneity in infectious disease emergence probability; however, how this information can be leveraged to optimize sentinel node selection for early outbreak detection on complex networks remains largely unexplored.

**Methods:** We simulated outbreaks on diverse synthetic and empirical networks, and quantified early detection performance as the average reduction in outbreak size when detected by a given sentinel set. We first used genetic algorithms to identify optimal sentinel sets and understand characteristics potentially relevant to early detection performance. We then trained a Random Forest-based Surrogate Model (RFSM) with the identified characteristics to assess the relative importance of different network features and to enable a rapid prediction of node selection rank. RFSM was benchmarked against five alternative surveillance strategies on networks not used in training to evaluate generalizability. Sensitivity analyses were conducted to examine how feature importance varied with network structure and epidemiological parameters.

**Results:** Surveillance strategies incorporating emergence probability outperformed those based solely on network topology, but the improvement was modest across all examined scenarios. Dynamic selection features capturing overlapping information among sentinel sites, such as the proportion of a candidate node’s neighbors that have already been selected, were the most important determinant of early detection, followed by global and node topology-related features. Emergence probability-related features were less influential but gained importance with greater node degree heterogeneity, larger variability in the emergence probability distribution, and greater negative correlation between node degree and emergence probability. Selecting only six sentinels achieved approximately 90% of the performance of full-network surveillance. RFSM achieved comparable performance to Genetic Algorithm (GA), while requiring only 1/24,000 of GA’s computational time on a network with 200 nodes.

**Conclusions:** Information about spatial heterogeneity in emergence probability provided limited additional benefit beyond network topology in selecting sentinel nodes for early outbreak detection on complex networks. The online tool RFSM offers a ready-to-use, computationally efficient and robust framework to support the design of effective disease surveillance networks.

## Introduction

In recent decades, the frequency, geographic spread, and impact of emerging infectious diseases (EIDs) have escalated notably, driven by factors such as climate change, urbanization, and ecological disruption [1–3]. Since the beginning of the 21st century, more than 50 EIDs have emerged, with at least one EID appearing nearly every year [2], some leading to large outbreaks. Beyond the direct health impacts, EIDs have also resulted in substantial socioeconomic burdens, including increased healthcare costs, disrupted trade, and widespread societal consequences. For example, the COVID-19 pandemic resulted in $52 billion in healthcare costs in low- and middle-income countries in the first 4 weeks alone [4], and led to a 3.4% decline in the world’s collective gross domestic product (GDP) in 2020 [5]. These challenges highlight the need for improved early detection of EIDs to strengthen global health security.

Early detection of EIDs has been addressed by researchers from two main perspectives. The first focuses on the spatial heterogeneity of disease emergence risks [6]. EIDs most frequently occur in remote, ecologically diverse regions (e.g., forests and wetlands) characterized by high wildlife density and low human population density, since most EIDs originate from zoonotic spillover events before its persistent transmission in humans, with over 60% arising from wildlife reservoirs such as bats, rodents, and primates [1]. Occasionally, these localized transmissions further spill over into densely connected urban centers, fueling larger outbreaks [7]. For example, human immunodeficiency virus (HIV) emerged from cross-species transmission of simian immunodeficiency viruses (SIVs) in African primates, spread to urban populations in Central Africa, and was later identified in multiple regions worldwide in the late 20th century [8,9]. Considerable efforts have been made to understand the geographic distribution of the emergence probability and identify disease emergence hotspots. The PREDICT project of the United States Agency for International Development’s Emerging Pandemic Threat (EPT) program conducted extensive wildlife sampling in regions with high zoonotic pathogen diversity and frequent human-animal contacts to enhance global surveillance and preparedness for EIDs [10]. Similarly, modeling studies have been developed to predict global hotspots and determinants of zoonotic disease emergence based on historical EID events [1,6,11]. However, how this information can be leveraged to optimize sentinel site selection for early outbreak detection on complex networks (i.e., neither purely regular nor random in structure) remains largely unexplored.

The other perspective emphasizes the critical role of network topology. Highly connected individuals or locations within a network are often infected earlier, thus offering timely signals of epidemic outbreaks [12,13]. In scale-free networks, for instance, monitoring highest-degree nodes can predict outbreak peaking time earlier than monitoring randomly selected nodes, reducing the peak detection time by up to 85.7% relative to random monitoring [12].

Beyond node degree, other network metrics—such as the degree of neighboring nodes [14,15] and the entropy of random walk from node to each community in networks with distinct community structure [16]—have also been leveraged to enhance EID surveillance system design, since they require less information of the complete network structure and remain robust to changes in network topology or missing information. However, these network-based studies typically assume uniform disease emergence risk across all nodes and rarely account for the spatial heterogeneity of such risks.

In this study, we integrated these two key perspectives—spatial heterogeneity of disease emergence risks and network topology—to identify the optimal set of sentinel nodes for early detection of EIDs in complex networks. By simulating outbreaks on diverse synthetic and empirical networks under different node-specific emergence probability scenarios, we evaluated the potential benefits of incorporating heterogenous emergence probability, proposed a computationally efficient method to identify optimal nodes without simulation, and examined how surveillance performance changes with the number of sentinel nodes and network size. We also developed an online platform implementing this analytical framework to facilitate evidence-based surveillance system design. This framework is also applicable to non-endemic but outbreak-prone infectious diseases.

## Methods

### Simulation settings

#### Network structures

We simulated infectious disease spreads on undirected and unweighted graphs *G* = (*V*, *E*), where *v_i_* ∈ `V` represents a vertex (also known as a node) such as an individual, a farm, or a city, and (*v_i_*, *v_j_*) ∈ `E` represents contacts between vertices, such as co-location in physical space, livestock exchange, or direct flight. The degree of a vertex is defined as the number of edges attached to it. We used synthetic modular networks for the main analyses due to its flexibility in reflecting diverse real-world network structures through changing key parameters [13], while further validated the results on simpler synthetic scale-free networks [17] and four empirical networks reflecting actual patterns of interaction and social structure [18–22].

##### Synthetic modular networks

Modular networks are ubiquitous in real-world, since they reflect the naturally occurring groups or community structure in society [13,23]. In such networks, connections are dense within groups (i.e., modules or communities), while relatively sparse between groups. We used Modular Configuration Model with predefined mean degree (μ), variance of the degree (σ), and within-module connection probability (*p*) (see instances in Fig. 1A-i) to generate synthetic modular networks [13,24]. We drew 1,000 sets of these parameters from predefined ranges (Table 1) to generate random networks and simulate disease outbreaks on them, ensuring sufficient diversity in the training data for the *Random forest-based surrogate model*.

**Figure 1.**
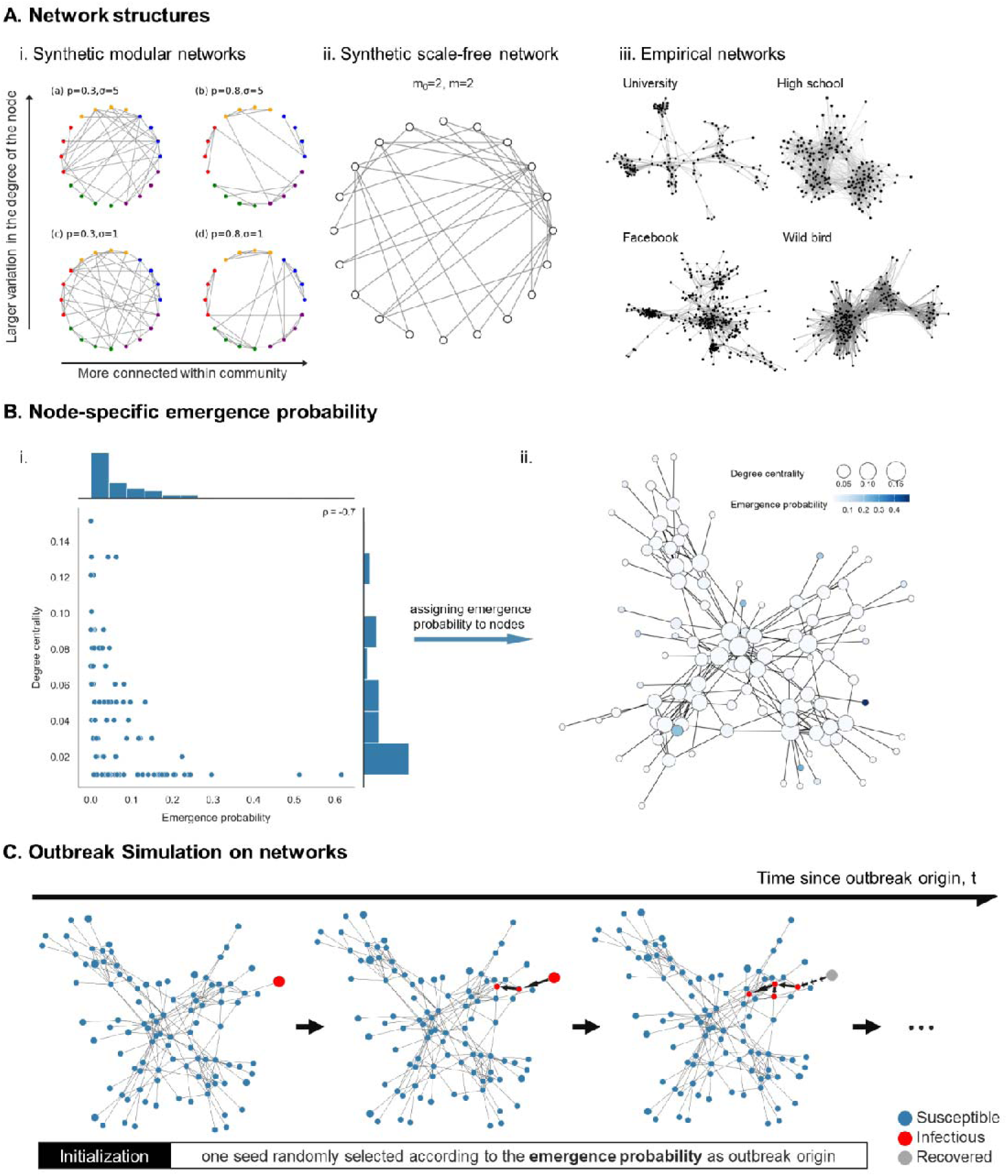
Simulation settings. (A) Example synthetic and empirical networks. (i) Four representative networks with 20 nodes (5 modules, 4 nodes per module) generated with the Modular Configuration Model, where *p* denotes the within-module connection probability and σ represents degree heterogeneity (Table 1). Colors distinguish different modules. (ii) A random synthetic scale-free network with 20 nodes generated with the Barabási-Albert model. (iii) Four empirical networks: human contact networks including university, high school, and Facebook social networks, and an ecological interaction network of wild birds. (B) Node-specific emergence probability assignment. (i) An example scatterplot of the emergence probability and node degree centrality when *N_m_*= 5, *N_n_* = 20, μ = 5, σ *=* 8*, p =* 0.8, *a* = 0.1, *b* = 5, and ρ = -0.7. (ii) An example synthetic modular network, whose node size represents degree centrality and node color represents emergence probability. (C) Simulated infectious disease spread on a modular synthetic network with the same parameters as above using a stochastic SIR (Susceptible-Infectious-Recovered) model. The outbreak was seeded from a node randomly selected based on its emergence probability.

**Table 1.**
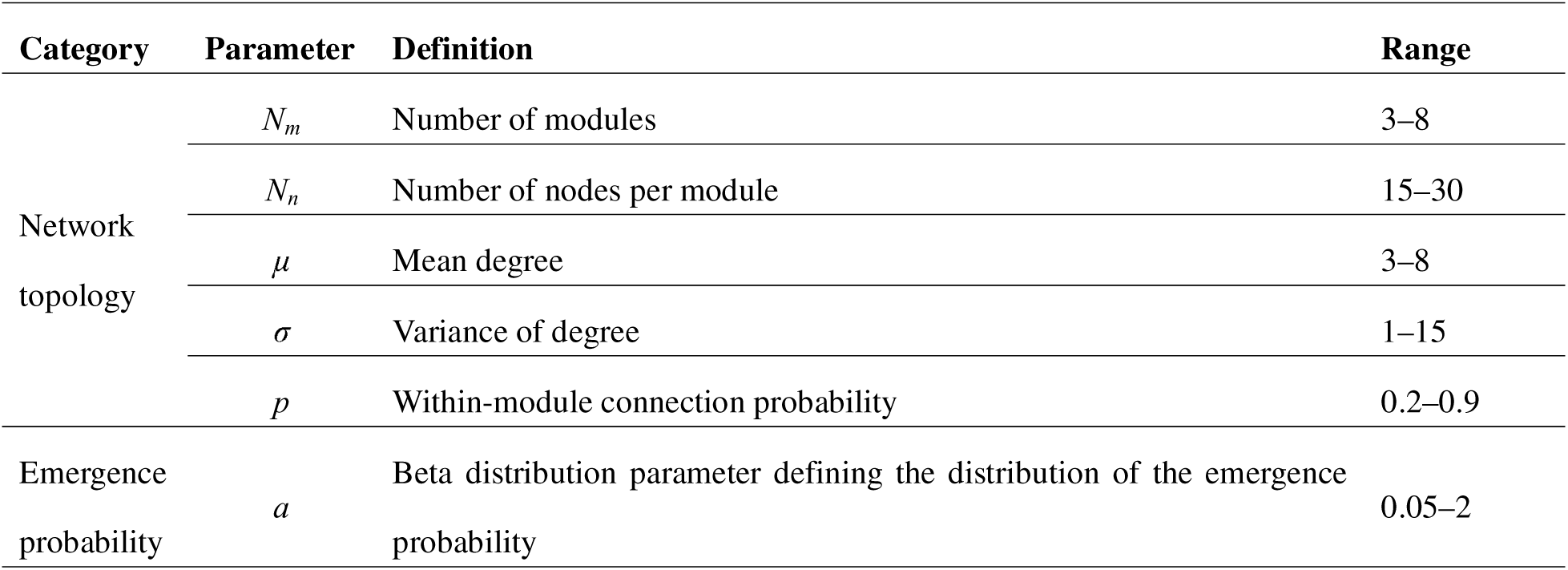
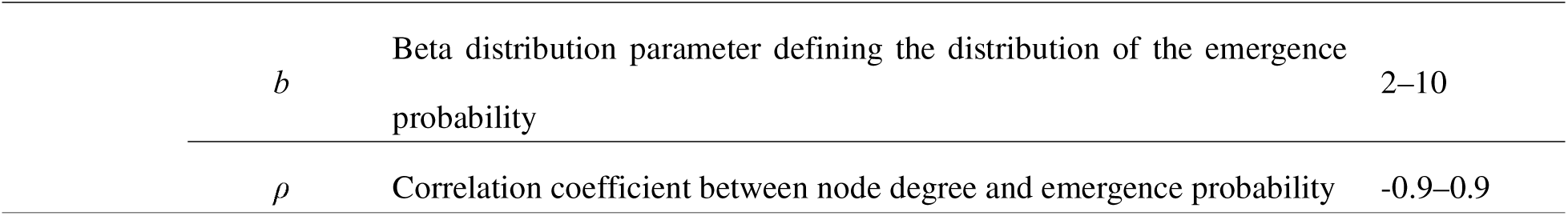
Parameter values related to network topology and emergence probability used to generate simulated networks and outbreak scenarios.

##### Synthetic scale-free networks

Scale-free networks, characterized by the existence of highly connected hubs, are another typical type of complex networks [17]. The degree distribution of these networks follows a power law, which means that most nodes have only a small number of connections, while a few nodes (i.e., hubs) have a large number of connections [17]. We generated synthetic scale-free networks with 100 nodes with the Barabási-Albert model to validate our surrogate model [17,25], beginning with an initial set of 2 nodes and iteratively adding new nodes, each connecting randomly to 2 existing nodes with the connection probability proportional to each existing node degree (Fig. 1A-ii).

##### Empirical networks

We used four medium-sized empirical networks to validate our findings (Fig. 1A-iii), chosen primarily for computational feasibility. Since most of empirical networks are not fully connected, we restricted our analyses to the largest connected components of each network.

- *University network.* The university network was collected via smartphones as part of the Copenhagen Network Study and contains information about physical contacts between university students defined as the reception of mutual Bluetooth scans between two students [18]. Since the original dataset is dynamic (i.e., every 5 minutes over 4 weeks) and using aggregated data across all time points will contain too many edges and obscure important network structures, we chose data from the time point *t* = 139,800s when student users were most active, resulting in a network with 143 nodes and 635 edges.
- *High school network.* This network captures physical contacts recorded by wearable sensors between students at a high school in Marseille, France during a 7-day period in November 2012, comprising 180 nodes and 2,220 edges [19].
- *Facebook network.* This dataset comprises an anonymous Facebook friends network collected in Fall 2014, containing 362 nodes (each representing a unique Facebook profile) and 1,988 edges (denoting Facebook friendship connection between profiles) [20].
- *Wild bird interaction network.* This network was derived from a socio-ecological study of wild birds at Witham Woods, Oxford, UK, conducted over a 24-day period in March 2014 [21,22]. It contains 202 nodes and 4,574 edges, where nodes represent wild birds and edges represent simultaneous presence of two birds at the same nest box.

The degree distributions of these empirical networks show different structural patterns (Fig. S1). The Facebook network exhibits degree distributions consistent with scale-free networks, while university and high school networks show more homogeneous degree distribution across nodes. Notably, the wild bird network shows a bimodal degree distribution, probably because some birds form larger groups for breeding and brood rearing during the breeding season [21].

#### Node-specific emergence probability assignment

We first randomly sampled *N* emergence probabilities from a predefined distribution, then assigned these probabilities to nodes based on a predefined correlation between node degree and emergence probability (Table 1) with the process described below.

##### Sampling emergence probabilities

We assumed that the emergence probabilities of all nodes follow a Beta distribution, chosen for its flexibility in approximating common distributions, such as uniform, exponential, gamma, and normal distributions, through changing its parameters. We randomly sampled *N* emergence probabilities {*p*_1_, *p*_2_, …, *p_N_*} from Beta (*a*, *b*), where *N* is the number of nodes, and *a* and *b* are the parameters of Beta distribution. The ranges for *a* and *b* are provided in Table 1.

##### Assigning emergence probabilities to nodes

We then assigned the sampled emergence probabilities to nodes using the method described below to achieve the predefined correlation coefficient ρ between node degree and emergence probability. Various correlation strengths were examined to compare the relative importance of emergence probability and network topology (Table 1). Under a strong positive correlation scenario, it is intuitively that nodes with both high emergence probabilities and large degrees will be prioritized.While under a strong negative correlation scenario, nodes with high degrees tend to have low emergence probabilities, and vice versa. In this setting, the prioritized characteristic (i.e., network topology or emergence probability) directly demonstrates its importance for early detection performance. To achieve the predefined correlation strength ρ, we first sampled *N* data points *Q* = {(*x*_1_, *y*_1_), (*x*_2_, *y*_2_), …, (*x_N_*, *y_N_*)} from a standard bivariate normal distribution with a correlation coefficient ρ. Then, for each node *V_i_*with degree *D_i_*, we determined its rank *R*(*D_i_*) relative to all other nodes’ degrees and identified the data point (*x_t_*, *y_t_*) in *Q* where the rank of *x_t_* matched *R*(*D_i_*), and determined the rank of *y_t_*, *R*(*y_t_*) among all *ys* in *Q*. Finally, the emergence probability with the rank *R*(*y_t_*) was assigned to *V_i_*. Ranks for nodes with identical degrees (e.g., {*V_i_*_1_, *V_i_*_2_, …, *V_i_*_s_}) were assigned based on their original order in the node list (*i*_1_ < *i*_2_ < … < *i_s_*) to ensure distinct emergence probabilities. Since the original node order was randomly assigned, this arbitrary rank allocation scheme should not bias the results.

#### Outbreak simulation on networks

We simulated 1,000 random disease outbreaks with a stochastic Susceptible-Infectious-Recovered (SIR) process on each synthetic or empirical network. For each simulation, we first sampled and assigned an emergence probability to each node, following the method described above. Then, we initialized the state of all nodes to S (susceptible) and randomly selected a seed node by sampling with probability proportional to its emergence probability, set its state to I (infectious), and simulated the onward transmission from this seed node (Fig. 1C). During each step, infectious nodes independently attempted to infect their susceptible neighbors with a transmission rate β, and transitioned to R (recovered) after an infectious duration y. We fixed y at 1 following previous studies [13,26] and calculated β to achieve target R_0_ values using the relationship β = R_O_/((< k^2^ > -< k >)⁄<k >), where <k > and < k^2^ > are the mean degree and the mean squared degree, respectively [27]. To explore varying transmissibility on the results, we used R_0_=3.0 for the main analyses to represent a moderately high transmission scenario that ensures sufficient epidemic spread for robust comparison across surveillance strategies. Sensitivity analyses were conducted using R_0_=1.5, 2.0, 2.5.

### Surveillance performance assessment

The surveillance performance of a sentinel network was evaluated as its ability to enable early detection of ongoing infectious disease transmission following previous studies [13,28–30]. Formally, given a predefined number of sentinel nodes *l*, we seek a subset *S* = {θ_1_, θ_2_, …, θ*_l_*} of nodes in the network that maximize the objective function Φ(*S*), which was defined as the average difference between the full outbreak size and the outbreak size at the time of detection (i.e., when the outbreak first reaches any node in *S*) across 1,000 simulated disease outbreaks.

Conceptually, Φ(*S*) can be treated as the average number of cases that can be prevented by *S* when timely control measures are implemented after detection. Mathematically, Φ(*S*) can be expressed as:

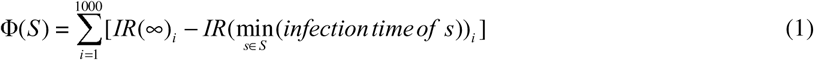

where *IR*(*t*) is the total number of nodes in the states I and R at time *t*. We additionally defined Φ(*S_all_*) as the objective function value when all nodes were included in *S*, representing the ideal surveillance performance under unlimited resources and serves as a benchmark for quantifying the relative performance of a given surveillance design. The impact of sentinel network size *l* on surveillance performance was further examined in *Changes of early detection performance with the number of sentinel nodes*.

### Optimal sentinel node selection using a genetic algorithm

We first used a genetic algorithm (GA), widely applied in surveillance system optimization [31–34], to identify the optimal subset of *l* nodes in the network that maximizes the objective function Φ(*S*), as exhaustive enumeration is computationally infeasible in large networks. The results of GA provide a benchmark for evaluating less computationally intensive node selection strategies; also, the optimal sentinel subset identified by GA can be used to understand key node characteristics contributing to early detection performance.

The GA process began by randomly generating 100 sets of *l* nodes as the initial population and evaluating the objective function value (also referred to as *fitness* in the context of GA) for each set (referred to as *individual* in GA). Then, these initial 100 individuals were sampled with replacement with the sampling probability of each individual proportional to its fitness, and randomly matched to 50 pairs. Each pair then produced two offspring for the next generation, with crossover and mutation happened with probabilities of *p_crossover_*= 0.8 and *p_mutation_* = 0.05, respectively, consistent with previous studies [35–37]. We applied the uniform crossover method to recombine parent solutions [38]. Specifically, if the two parent individuals are *F*_1_ = {*F*_11_, *F*_12_, …, *F*_1*l*_} and *F*_2_ = {*F*_21_, *F*_22_, …, *F*_2*l*_}, where each *F_ij_* represent a node ID, the two offspring (*O*_1_ and *O*_2_) were generated by independently determining node inheritance at each position through random number generation. For each position *i* (1 ≤ *i* ≤ *l*), a random number was drawn from a uniform distribution between 0 and 1. If the number was less than 0.5, *O*_1_ inherited *F*_1*i*_ and *O*_2_ inherited *F*_2*i*_, otherwise the assignments were swapped. When duplicate nodes appeared in an offspring, they were removed and the gaps were filled by randomly selecting unused nodes from the network *G*. When mutation occurred, a randomly chosen node in the offspring was replaced with a randomly selected unused node from network *G*. This evolutionary process was repeated iteratively until both the fitness value and node composition kept stable over 30 consecutive generations.

### Random forest-based surrogate model

Since the optimization process is computationally intensive and time consuming, we attempted to develop less demanding surrogate models to facilitate sentinel nodes selection within a shorter time frame. By observing the optimal node sets selected by GA under various settings and drawing on previous literature [13,39–50], we identified 28 characteristics that may influence the surveillance performance of a given node set, which were further grouped into 6 categories (Table 2). We chose to use a random forest given its ability in learning complex nonlinear relationships and high-order interactions between predictors, resisting overfitting via ensembling and providing accessible measures for variable importance [51,52]. The model was trained based on simulation data generated from synthetic modular networks and validated on both synthetic scale-free and empirical networks. We also used the fitted model to quantify the relative importance of these characteristics in determining node surveillance performance (see *Key node characteristics determining early detection performance*).

**Table 2.**
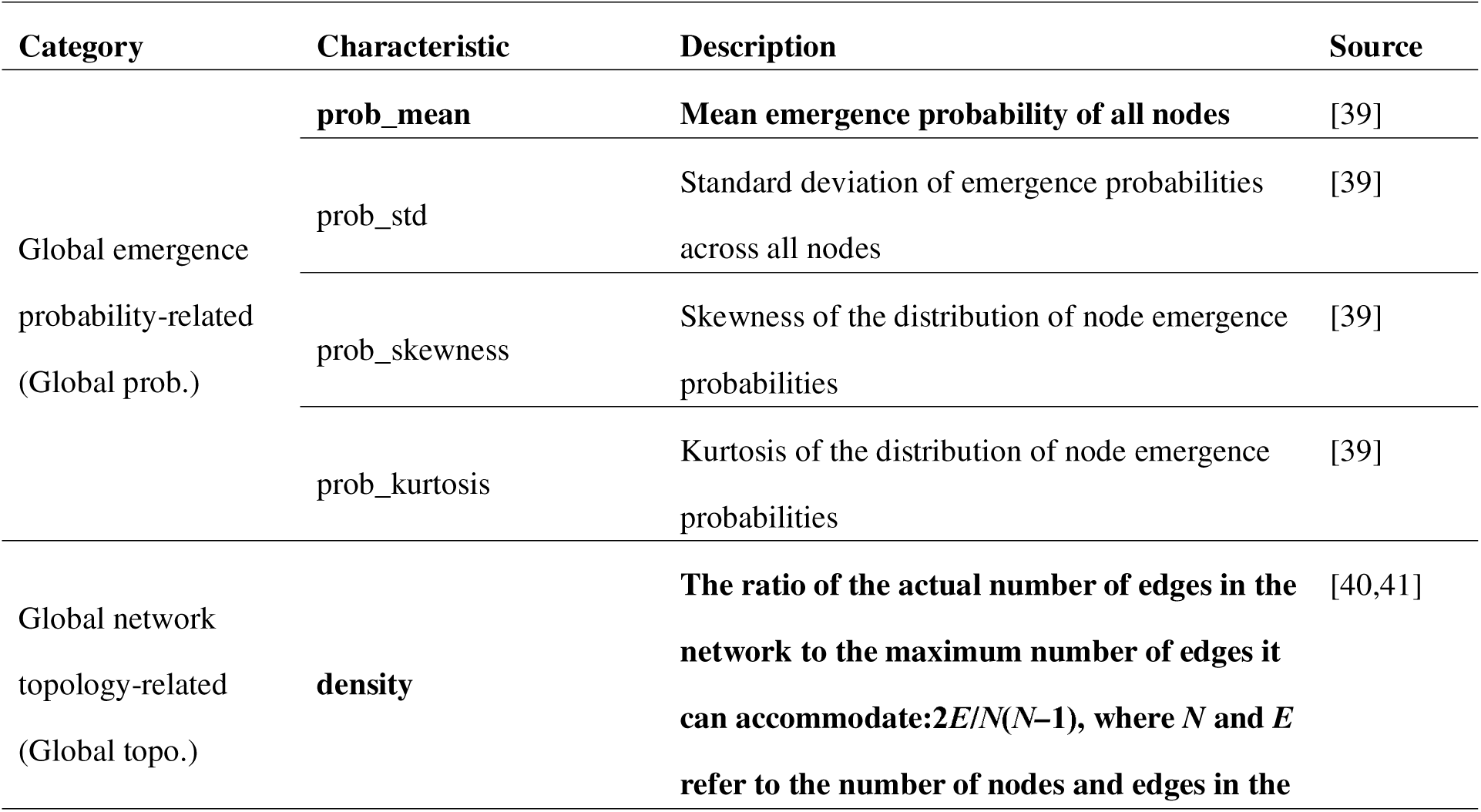
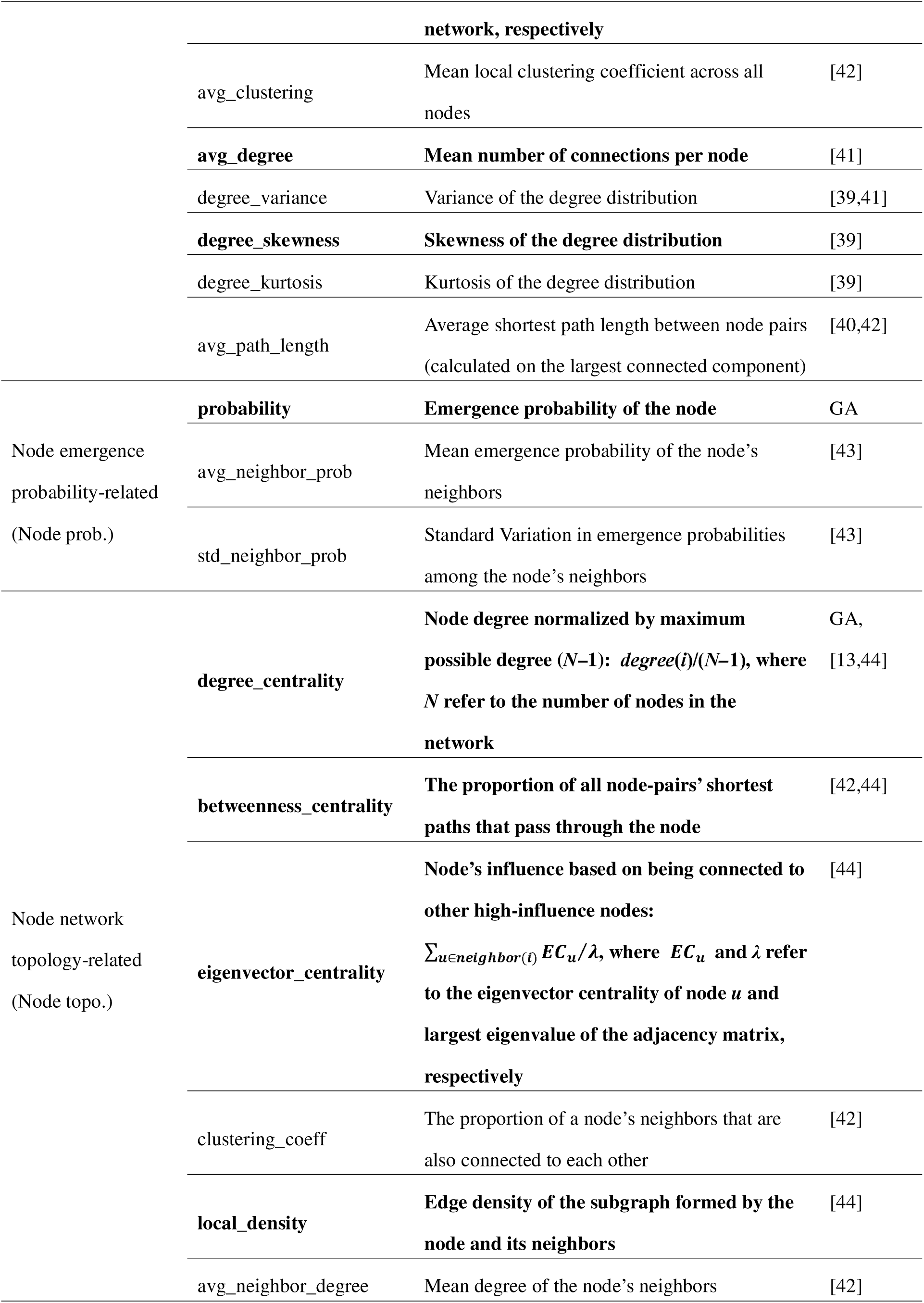
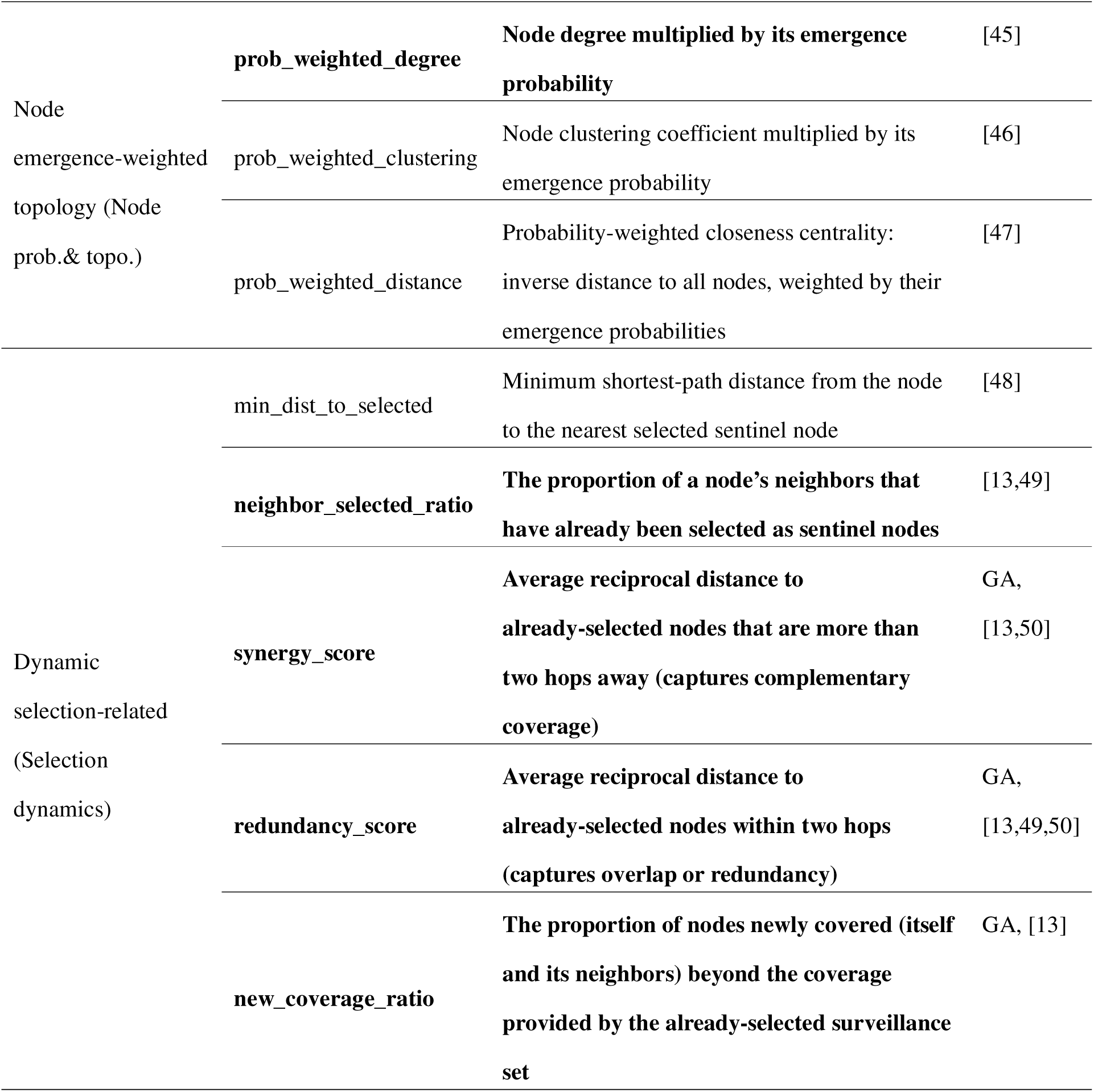
All candidate global- and node-specific characteristics considered in RFSM. Characteristics shown in bold indicate those selected in the final model.

#### Training data

We first generated 1,000 random modular networks using parameter sets randomly sampled from the ranges in Table 1 to ensure network structure diversity in the training data. To maintain sufficient complexity for meaningful pattern learning, we excluded four networks with fewer than 20 nodes or exhibiting extremely low emergence probability variance (σ_p_< 0.005). For each remaining network, we simulated 1,000 outbreaks and calculated the node characteristics listed in Table 2 as the independent variables. Near-optimal sentinel nodes were identified stepwise using a greedy algorithm that selects one node at a time iteratively from the remaining unmonitored nodes to maximize the marginal gain in Φ(S), and the relative selection order of each node was recorded as the dependent variable for model fitting (e.g., a node selected in the 3rd step was assigned a value of 3/N where N is the network size), since we found that the greedy algorithm performs comparably to the GA while being more computationally efficient (Fig. 5). We used relative rank order, rather than the surveillance performance value Φ(*S*), as the dependent variable, since in real-world applications the order of sentinel node selection will be used to guide sentinel site selection rather than the absolute value of Φ(*S*). In addition, the value of Φ(*S*) varied substantially with network size and *R*_0_, introducing unnecessary variability into model training. Only nodes ranked the top 30% from each network were used to fit the model, since the primary objective was to identify the most influential nodes rather than establishing a complete network-wide ordering. This process yielded a final dataset of 30,815 node-ranking pairs. We also repeated the analyses using the top 20% and 10% nodes, resulting in 20,800 and 10,663 node-ranking pairs, respectively.

#### Model fitting

We randomly split the dataset into a training (70%) and a testing set (30%) based on network membership, ensuring that all nodes from the same network were assigned to the same partition. A random forest model was fitted to the training set using all features listed in Table 2 as independent variables and the selection order derived from the greedy algorithm as the dependent variable. Hyperparameters, including the number of trees, maximum tree depth, minimum split and leaf sizes, feature sampling ratio, and bootstrap sample ratio, were tuned by Bayesian optimization with five-fold cross-validation to minimize the normalized discounted cumulative gain (NDCG@k) between observed and predicted ranks as the optimization metric [53]. NDCG was chosen over the commonly used mean squared error (MSE) because it directly measures the consistency between predicted and observed ranks, which better aligns with real-world applications, where the ordering of sentinel node selection matters more than the absolute magnitude of prediction errors. NDCG@k was calculated as:

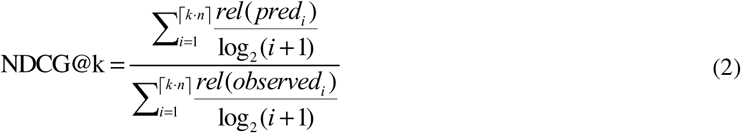

where *k* represents the percentage of top-ranking nodes considered in the performance measure, *n* represents the total number of nodes in the network, ⌈·⌉ is the ceiling function, *pred_i_* and *observed_i_*represent the *i*th node in the ranking lists produced by the model and the *greedy* algorithm, respectively, and *rel*(·) represents the relevance scoring function of a node, which refers to the inverse rank of the node within the observed list (e.g. the relevance score of the first node in the observed list *rel*(*observed*_1_) is *N* for a network with *N* nodes). NDCG@k ranges from 0 and 1, with 1 indicating that the predicted ranking matches the observed ranking. Permutation importance scores, defined as the decrease in NDCG after randomly permuting a given feature while keeping the other features unchanged, were obtained from the trained model with the optimal hyperparameters. Features with importance scores less than 0.01, a threshold value chosen based on a natural elbow in the importance ranking distribution (Fig. S2) that clearly distinguished influential features from other features, were excluded and hyperparameters were reoptimized on this reduced feature set using the same procedure. After evaluating the model performance on the testing dataset, the final model was refitted on the entire dataset with the optimal hyperparameters and used for further analyses.

#### Model validation

We externally validated the performance of RFSM on the unseen synthetic scale-free and empirical networks. For each network, node characteristics listed in Table 2 were computed as the inputs to the trained RFSM. Sentinel node sets were then defined based on the predicted selection ranks, and their performance was evaluated and compared against alternative surveillance strategies described below.

#### Benchmarking against alternative surveillance strategies

We evaluated the performance and computational efficiency of RFSM against five alternative surveillance strategies (Fig. 2), including selection of nodes with the highest degree in the whole network (*Global*), selection of nodes with the highest degree within their module (*Modular*, for synthetic scale-free and empirical networks, modules were partitioned by an improved version of the Louvain modularity algorithm [54] in Gephi [55]), random selection (*Random*), genetic algorithm-based selection (*GA*), and stepwise greedy selection (*Greedy*). These alternative strategies fall into two board categories: those relying solely on network topology without outbreak simulations (Figs. 2A–C, *Global*, *Modular* and *Random*) and those requiring outbreak simulations (Figs. 2E–F, *GA* and *Greedy*). Although the first group is computationally efficient, they may ignore nodes with high emergence probabilities or with high transmission capacity but moderate degree, such as bridge nodes with high betweenness centrality that connect different modules, or nodes embedded in high-risk neighborhoods that can accelerate secondary spread. In contrast, the second group, which involves outbreak simulations, can dynamically adjust sentinel node selection strategy by simulating the epidemic propagation process, enabling more accurate assessment of each node’s surveillance potential, but are at substantially higher computational cost. RFSM lies between these two categories as it requires simulations during model training by the developers, but not during applications by end users once the model is fitted. To ensure comparability, all strategies were benchmarked on identical network configurations and emergence probability settings.

**Figure 2.**
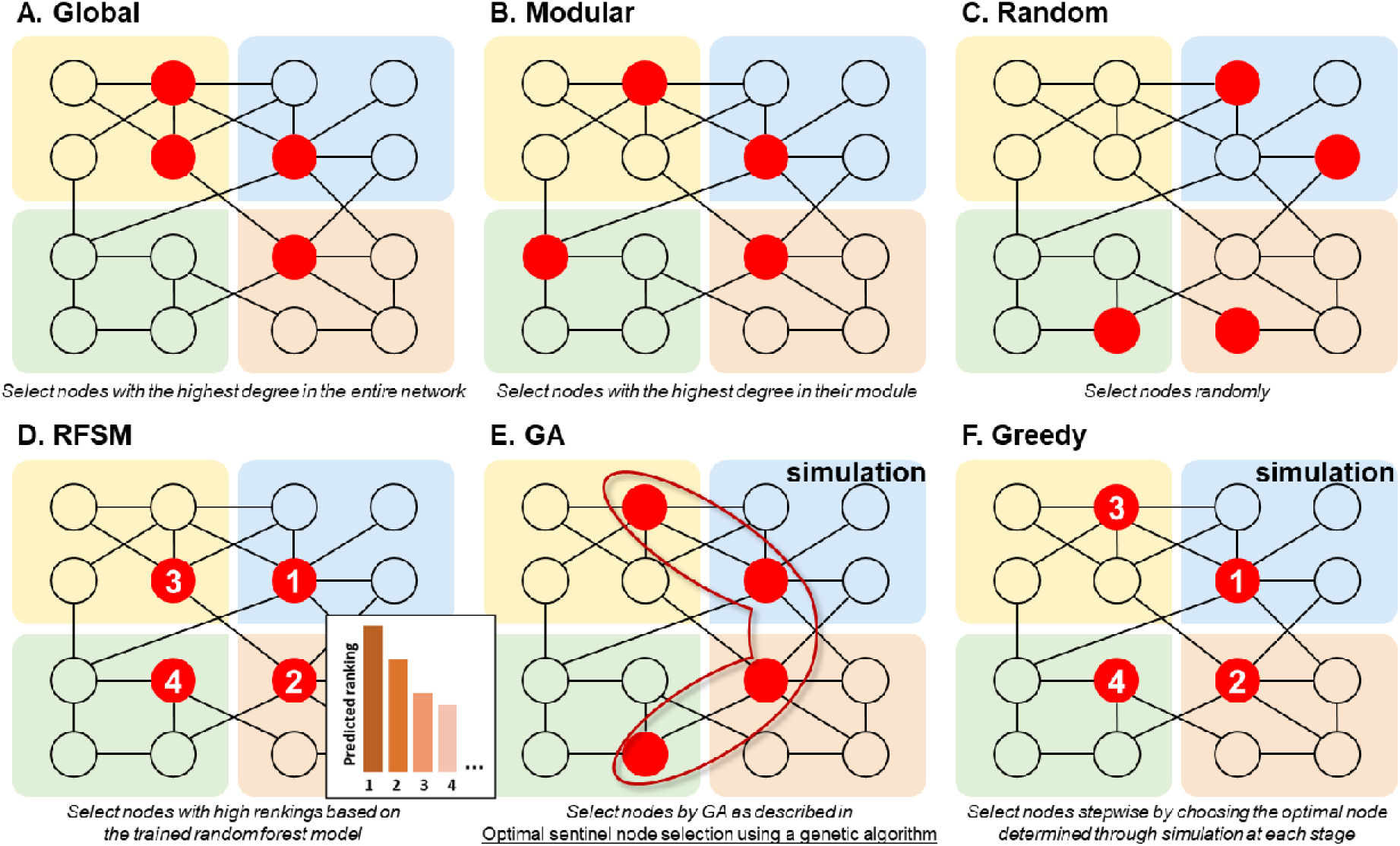
Schematic representation of six surveillance strategies. Red circles indicate nodes that are selected to be sentinel nodes, and colored backgrounds denote network modules.

#### Performance with incomplete network structure data

In real-world applications, the network structures are often only partially observed. To evaluate the robustness of the RFSM results to incomplete network structure observation, we randomly removed 10–90% of the nodes or edges from the complete network and applied RFSM to the resulting incomplete networks. Surveillance performance was evaluated on the original complete network prior to node (or edge) removal. The removal process was repeated 10 times to account for uncertainty in the removal process. Network and model parameters were set as *N_m_* = 5, *N_n_* = 20, μ = 5, σ = 8, *p* = 0.8, *a* = 0.1, *b* = 5, ρ = -0.7 and *R*_0_ = 3.0, as in the baseline scenario. We examined the scenarios when selecting 3, 6, or 9 sentinel nodes, since we found selecting only 6 sentinels can already achieve a high surveillance performance (Fig. 7).

#### Online platform for RFSM

To support real-world applications of RFSM, especially in resource- and research-limited regions, we further developed an online platform (https://lunawang.shinyapps.io/sentinel_nodes_selection/). After uploading the network structure and node-specific emergence probability and specify the desired number of sentinel nodes, the platform can provide the optimal sentinel node set selected by RFSM, as well as visualizes the selected nodes within the network to aid interpretation.

### Changes of early detection performance with the number of sentinel nodes

We further investigated how early detection performance evolved with the number of sentinel nodes across each surveillance strategy. Specifically, we began with a single sentinel node and gradually increased the number of sentinel nodes until further additions yielded negligible performance gains. This approach allowed us to identify an optimal threshold that minimize the number of sentinel nodes to conserve resources while maintaining reasonable surveillance performance. Additionally, we examined whether this optimal threshold varied with network structure and size by using diverse synthetic and empirical networks with varying topology and sizes.

### Key node characteristics determining early detection performance

*Overall feature and category importance.* We obtained the permutation importance score for each feature in the final model with the method described above. To further assess the relative contribution of network topology and emergence probabilities, we calculated the group importance of each characteristic category (Table 2) through simultaneously permuted all features within the category while keeping features in the other categories unchanged. The resulting decrease in NDCG was recorded at the permutation importance for the entire category.

#### Rank-specific feature and category importance

To understand how the relative contributions of node characteristics evolved across selection steps (i.e., which characteristics were prioritized when only few sites were chosen, and which became more important as the sentinel set expanded), we trained separate random forest models for each selection rank *k* (i.e., *k* = 1 corresponds to the first node selected in the stepwise process). In each model, nodes whose rank *k_i_ = k* were labeled as positive, nodes with *k_i_* > *k* as negative, while nodes with *k_i_* < *k* (i.e., nodes already chosen at earlier ranks) were excluded, ensuring that each model specifically learned to identify the next sentinel from the remaining pool. When *k* = 1, dynamic selection-related features were excluded because no nodes had yet been chosen; otherwise, all features were included. Permutation importance was then computed for each rank-specific model and normalized within each rank so that the scores summed to 1. We focused on the first ten sentinel nodes, since changes in the relative importance of characteristics became negligible thereafter (Fig. S8).

#### Sensitivity analysis of feature importance

To examine whether the relative contributions of node characteristics remained stable across diverse network structure and emergence probability distribution, we conducted sensitivity analyses through refitting models after stratifying the dataset into quintiles based on total number of nodes (*N*=*N_m_*×*N_n_*), mean degree (μ), variance of degree (σ), within-module connection probability (*p*), kurtosis of the emergence probability distribution (derived from parameters *a* and *b*, quantifying the shape and heterogeneity of assigned probabilities), and correlation coefficient between node degree and emergence probability (ρ). For each parameter, the quintile-based datasets were labeled G1–G5, with G1 representing the lowest and G5 the highest values. Thresholds used for stratification for each parameter are shown in Fig. S3. We also performed sensitivity analysis by varying the basic reproduction number (*R*_0_) used in the SIR simulations to assess the robustness of feature importance under varying epidemic transmissibility. While the main training dataset corresponded to *R*_0_ = 3.0, three additional datasets were generated under *R*_0_ = 1.5, 2.0, and 2.5 and the remaining network and probability parameters were also randomly sampled from the ranges listed in Table 1. These four datasets were labeled G1–G4 in ascending order of *R*_0_ (i.e., G1 = 1.5, G2 = 2.0, G3 = 2.5, and G4 = 3.0).

All the analyses in this study were conducted using Python 3.12.4 and R 4.4.1. In Python, we used packages networkx 3.3 [25] for generating and loading networks, EoN 1.2 [56] for disease simulation and surveillance performance assessment, scikit-learn 1.7.0 [57] for RFSM and permutation importance, pandas 2.2.2 [58] and numpy 2.0.1 [59] for data processing. Visualizations were primarily generated using matplotlib 3.9.1 [60] and seaborn 0.13.2 [61] in Python, and further refined using packages ggplot2 3.5.2 [62], cowplot 1.2.0 [63] and ggsci 4.0.0 [64] in R. All the code and data used in the analyses are publicly available at: https://github.com/qu-cheng/EID_optimization.

## Results

### Optimal sentinel nodes selected by GA

We examined the optimal sentinel node sets selected by GA across various synthetic networks to identify key node characteristics associated with early detection performance. For illustrative purposes, we only present results of *l* = 6 sentinel nodes for a modular network with *N_m_* = 5, *N_n_* = 20, μ = 5, σ *=* 8*, p =* 0.8, *a* = 0.1, and *b* = 5 (Figs. 3A and 3B) and a synthetic scale-free network (Figs. 3C and 3D), both with a correlation of ρ = -0.7 between node degree and emergence probability. When emergence probabilities were homogenous across all nodes, sentinel selection prioritized nodes with the highest degrees (Figs. 3A and 3C). By contrast, when emergence probabilities were heterogenous, the optimal set reflected a balance between node degree and emergence probability (Figs. 3B and 3D). For example, in the baseline network, the optimal sentinel set include the node with the highest emergence probability (Fig. 3B), while in the scale-free network, selected nodes include both the node with the highest emergence probability and its neighbor (Fig. 3D). Furthermore, optimal sentinel nodes were typically distributed across the network, reducing redundancy and maximizing surveillance coverage. Based on these findings, we included characteristics related to node centrality, emergence probability, and especially dynamic aspects such as redundancy and coverage into subsequent analyses (Table 2).

**Figure 3.**
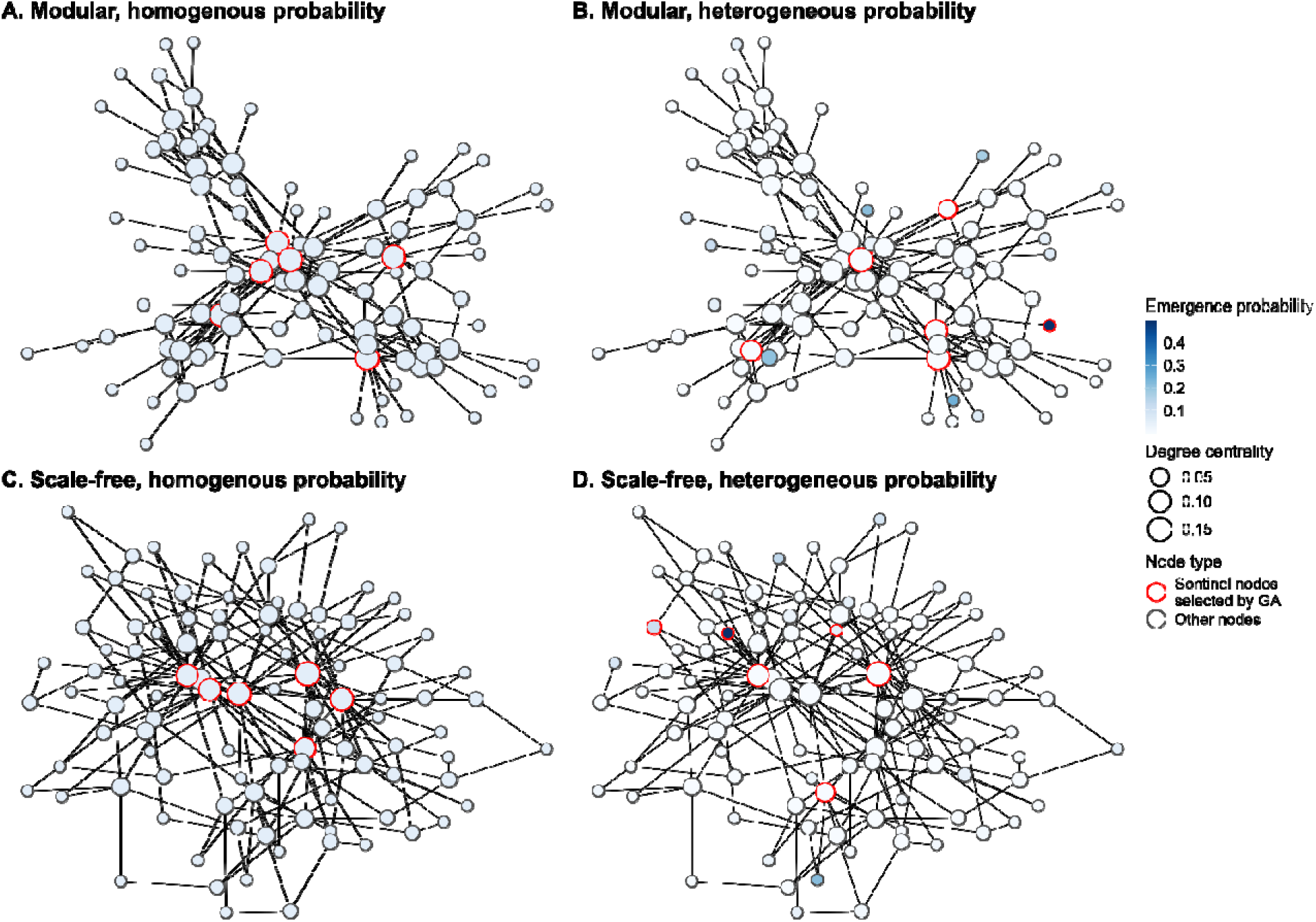
Optimal sentinel nodes selected by GA on synthetic networks. Comparison of GA-selected optimal sentinel nodes in (A) a modular network and (B) a scale-free network under homogenous emergence probabilities, and in (C) a modular network and (D) a scale-free network under heterogenous emergence probabilities. In (A) and (C), each node was assigned the same emergence probability of 0.1, while in (B) and (D), they were sampled from Beta (0.1, 5) with a correlation of ρ = -0.7 between node degree and emergence probability. The basic reproduction number *R*_0_ was set to 3.0. Node color and size indicate their emergence probability and degree, respectively. Optimal sentinel nodes selected by GA were highlighted in green for the homogenous scenario, while in red for the heterogenous scenario.

### Random forest-based surrogate model performance

During feature selection, 14 of the 28 features were retained for further analysis (highlighted in Table 2 in bold, Fig. S2). The final RFSM demonstrated strong predictive performance on both the training and testing datasets for the synthetic modular networks, achieving NDCG@30% values of 0.978 and 0.954, respectively (Fig. 4A). Moreover, the model also performed well on various validation networks, with NDCG@30% ranging between 0.943 for the university network to 0.996 for the high school network. These consistently high values underscore the model’s robust predictive capability on unseen data, even across networks with entirely different topological patterns.

**Figure 4.**
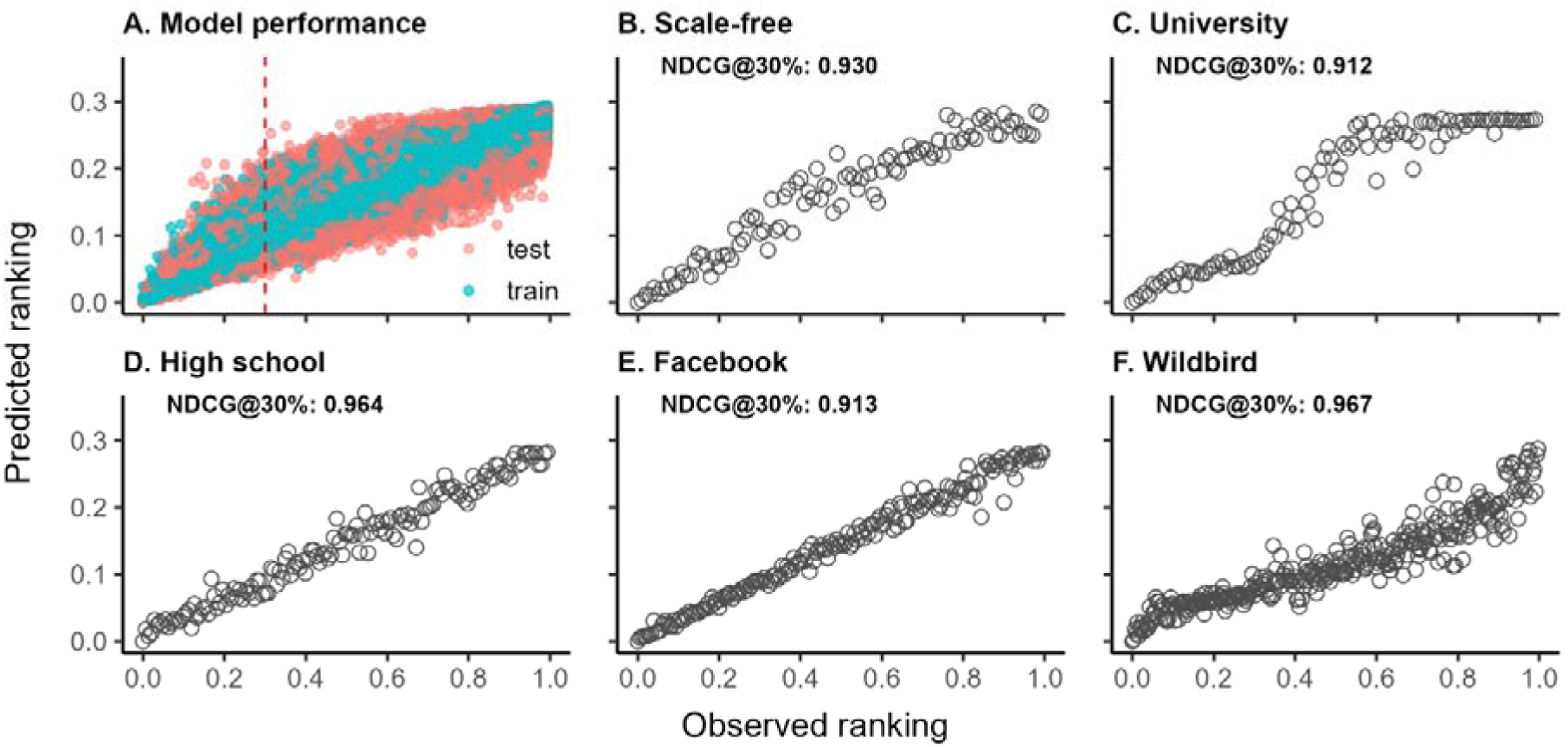
Performance of the final RFSM. (A) Model performance on the training and test datasets (vertical dashed line indicates the top 30%). (B–F) Evaluation of the trained RFSM across five networks: (B) synthetic scale-free, (C) university, (D) high school, (E) Facebook, and (F) wild bird contact networks. Each panel shows the relationship between observed and predicted node rankings, with model accuracy summarized by the normalized discounted cumulative gain (NDCG@30%) metric.

**Figure 5.**
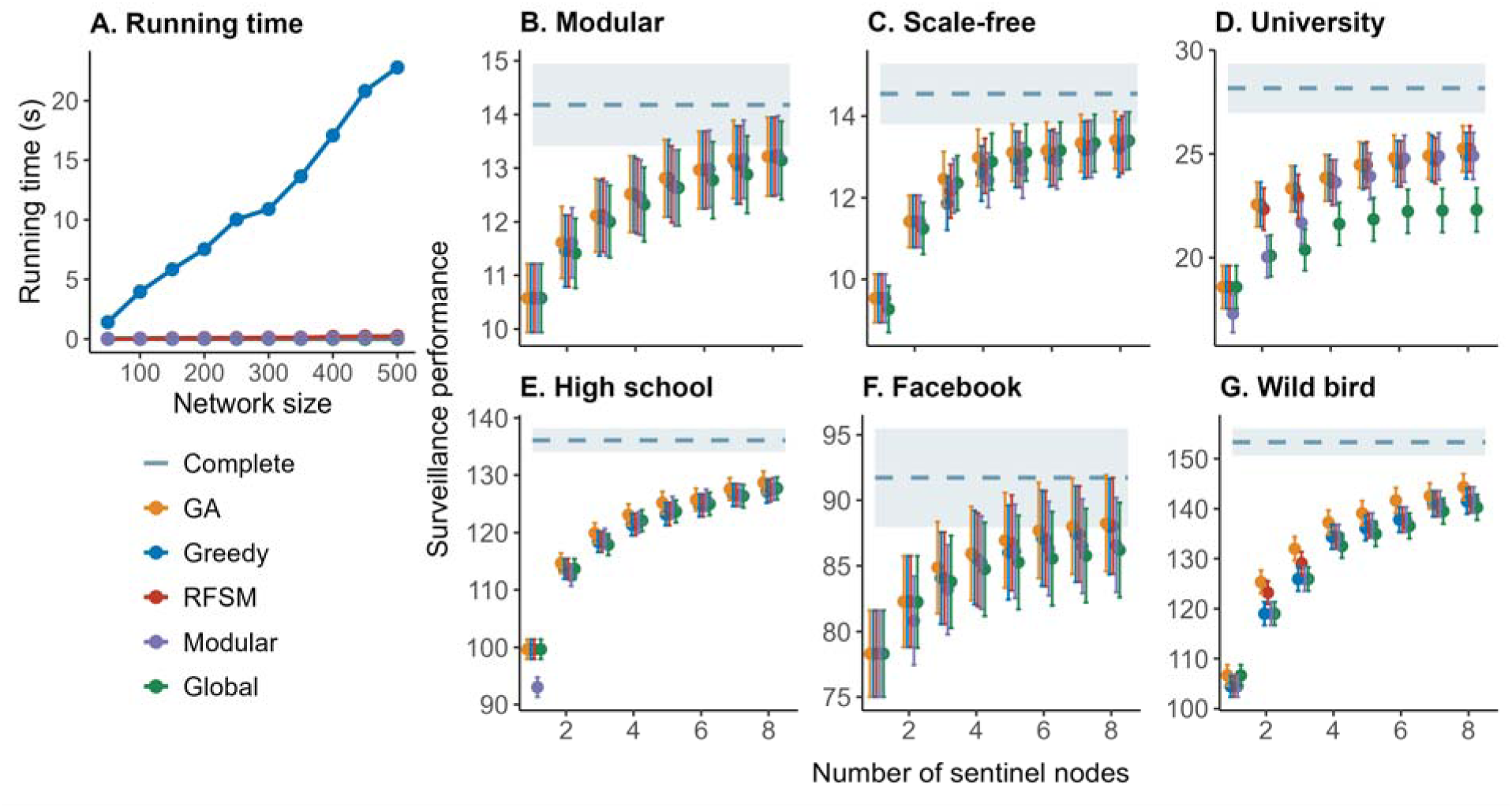
Surveillance and computational performance comparison across different networks. (A) Runtime (y-axis) is plotted against network size (x-axis) for RFSM, *Greedy*, *Global* and *Modular*. All synthetic networks maintain same topological properties (*N_m_* = 5, *N_n_* = 20, μ = 5, σ = 8, and *p* = 0.8) to the modular network with only node count varying from 50 to 500 and all measurements represent average runtime over 100 runs for selecting 6 sentinel nodes. (B–G) Each panel shows the surveillance performance (y-axis, defined as the number of infected nodes prevented) as a function of the number of sentinel nodes (x-axis) for different node selection strategies. The blue dashed line represents the surveillance performance of monitoring all nodes in networks. Error bars indicate standard deviation across 100 simulation runs. For each network, node emergence probabilities were sampled from Beta (0.1, 5) distribution, with correlation coefficient (ρ) of -0.7 between node degree and emergence probability, and basic reproduction number (*R*_0_) set to 3.0.

### Benchmarking RFSM against alternative surveillance strategies

When compared with alternative sentinel site selection strategies across diverse synthetic and empirical networks, the GA consistently has the highest early detection performance, followed by the *greedy* algorithm. The RFSM demonstrated comparable performance relative to *greedy*, and outperformed the network structure-based approaches, including g*lobal*, *modular,* and *random* selection, especially on the empirical networks (Figs. 5 and S4 for *random*). While achieving near-optimal surveillance performance across diverse network topologies, RFSM’s most notable advantage lies in its exceptional computational efficiency (Fig. 5A), which scales nearly linearly with network size. It required less than 0.08 seconds for networks with 200 nodes, compared with 8 seconds for *greedy* algorithm and 1,965 seconds for the *GA* for the same network on a high-performance cluster node with 256 GB of RAM and two 28-core 2.7 GHz Intel Xeon Gold 6258R processors (Supported by High Performance Computing Platform of Huazhong University of Science and Technology). The network structure-based methods demonstrated superior computational efficiency with near-constant runtime, but their performance tend to be lower than RFSM.

Although the surveillance performance of RFSM decreased with increasing proportion of node and edge removal, as expected, it remained comparable to that of GA, *global*, *greedy*, and *modular* strategies, and consistently outperformed the *random* strategy (Fig. 6). Similar patterns were observed when selecting 3 or 9 sentinel nodes (Figs. S5–6).

**Figure 6.**
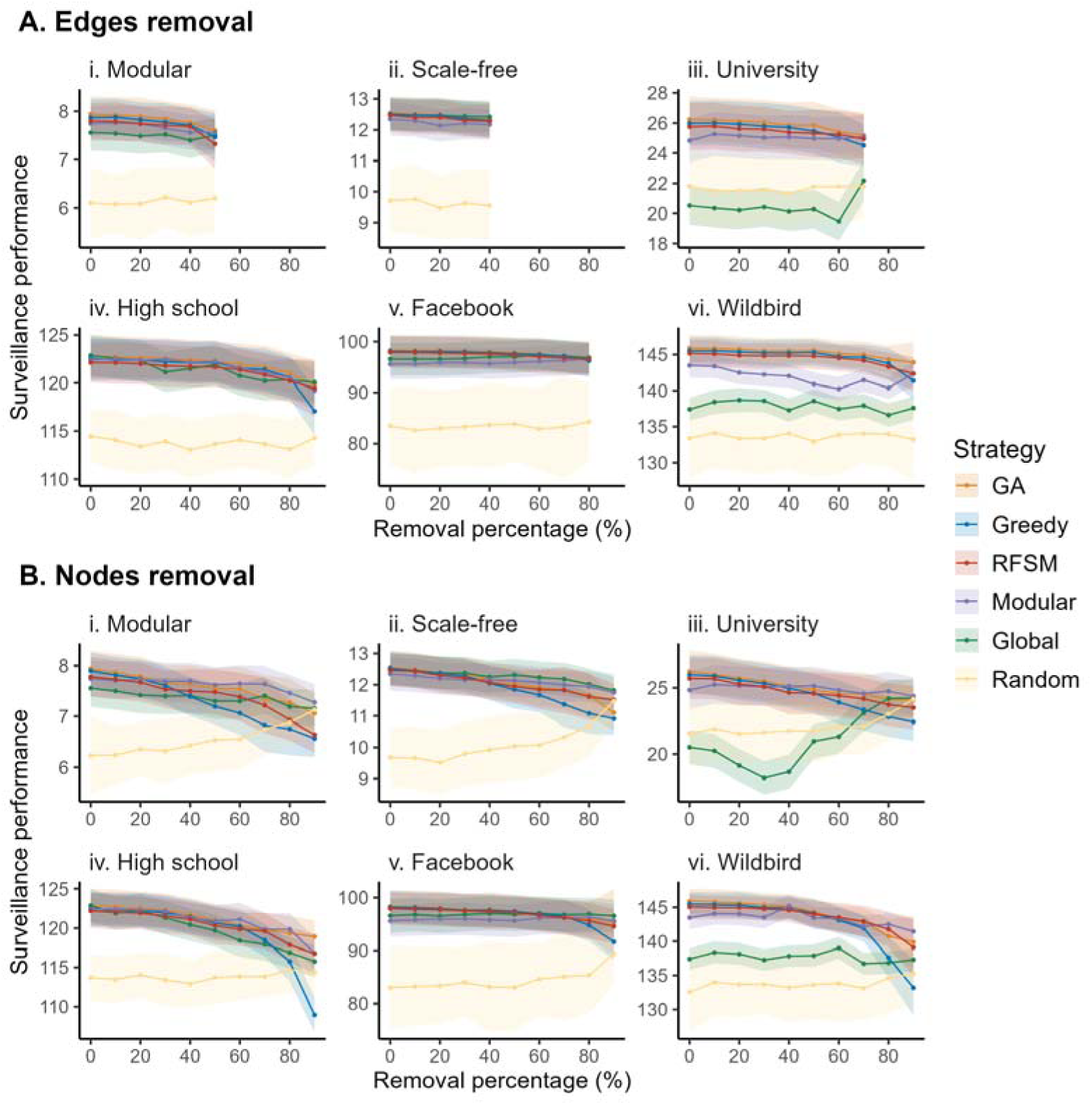
Surveillance performance under incomplete network structure observation when 6 sentinel nodes were selected. (A) shows results for edge removal, while (B) shows results for node removal. Columns (i–vi) correspond to different network types: Modular, Scale-free, University, High school, Facebook, and Wildbird. For each panel, the x-axis denotes the proportion of removed edges or nodes, and the y-axis represents mean surveillance performance, defined as the number of infected nodes prevented. Solid lines indicate average performance across simulations, and shaded areas represent variability across 10 repetitions.

**Figure 7.**
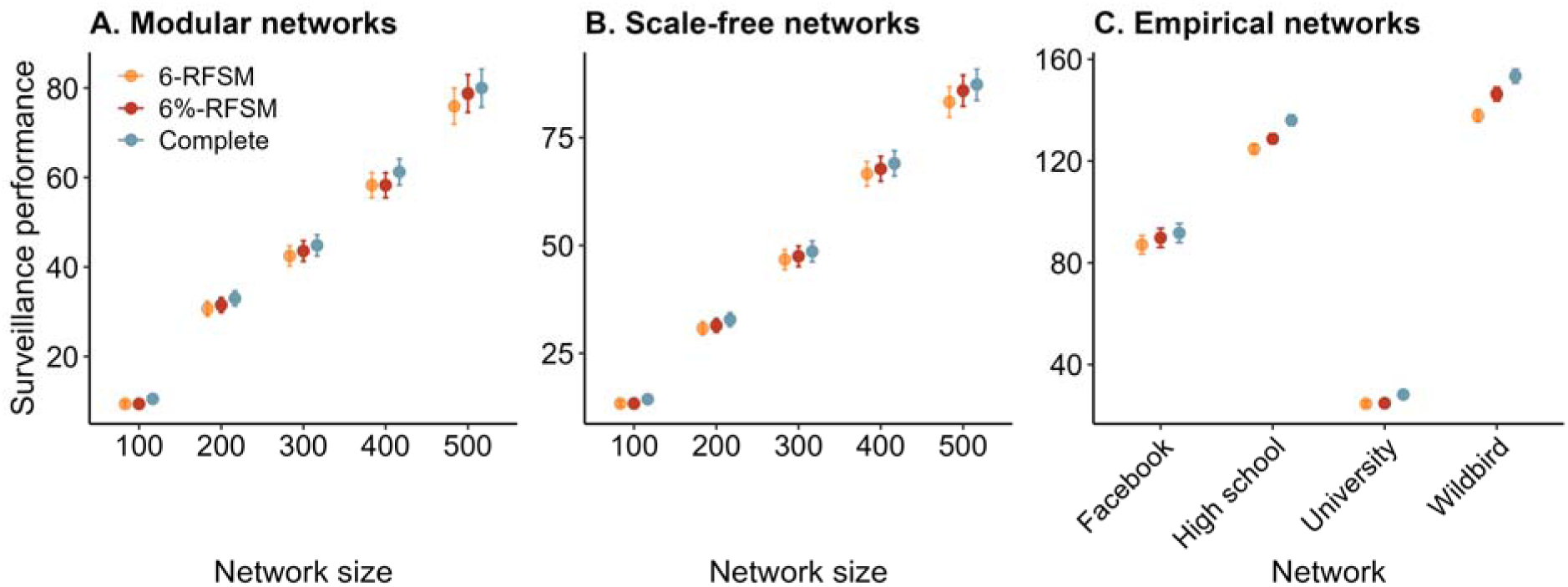
Surveillance performance comparison across network scales and types. Surveillance performance comparison across three monitoring scenarios: complete surveillance where all nodes are monitored, targeted surveillance using 6 RFSM-selected sentinels, and targeted surveillance using 6% of RFSM-selected sentinels. Performance comparisons are shown for (A) modular networks of different sizes, (B) scale-free networks of different sizes, and (C) four empirical networks (university, high school, Facebook, and wild bird contact networks). Surveillance performance (y-axis) represents the number of infected nodes prevented through early detection by the selected sentinel nodes. Each data point represents mean performance over 100 simulation runs, with error bars indicating standard deviation. For each network, node emergence probabilities were sampled from Beta (0.1, 5) distribution, with correlation coefficient (ρ) of -0.7 between node degree and emergence probability, and *R*_0_ set to 3.0.

### Changes of early detection performance with the number of sentinel nodes

Across all examined synthetic and empirical networks, the surveillance performance of all strategies improved with the number of sentinel nodes increased, though the marginal gain diminished as the sentinel set grew, especially beyond 6 sentinel nodes, where additional nodes yielded minimal improvement (Fig. 5B–G). To assess whether this optimal number of sentinel nodes scales with network size, we generated modular and scale-free networks of varying sizes and compared surveillance performance when all nodes, 6 nodes, or 6% of nodes were included as sentinels (Fig. 7). Results showed that while monitoring all nodes provided the best performance, using just 6 sentinel nodes offered substantial efficiency, performing comparable to monitoring 6% of nodes, suggesting that a small set of strategically chosen sentinels can achieve high surveillance performance without requiring proportional increase in the number of nodes.

### Key node characteristics determining early detection performance

#### Overall feature and category importance

Feature importance analysis of the retained features highlighted dynamic selection features as the dominant determinants of early detection performance (Fig. 8B), among which the proportion of a node’s neighbors already chosen as sentinels (*neighbor_selected_ratio*) was the most influential, showing a negative effect that nodes with many sentinel neighbors were less likely to be chosen (Fig. S7), reflecting the model’s tendency to minimize redundancy in coverage. Other important dynamic features included the average reciprocal distance to already-selected nodes beyond two hops away (*synergy_score*) and the proportion newly covered nodes (*new_coverage_ratio*), both suggesting a preference for nodes that extend cooperative coverage and extend monitoring into previously unobserved regions. Global network topology-related features ranked second in importance (Fig. 8B), with network density (*density*) being the most important variable in this category (Fig. 8A), highlighting the role of overall network connectivity in shaping sentinel selection dynamics. Node network topology-related features are the third most important (Fig. 8B), with centrality and local connectivity features (e.g., degree centrality *degree_centrality*, betweenness centrality *betweenness_centrality*, eigenvector centrality *eigenvecgtor_centrality*) providing modest improvements in predictive accuracy (Fig. 8A). In contrast, features related to emergence probability, both direct and topology weighted, contributed relatively little to the predicted surveillance performance (Fig. 8).

**Figure 8.**
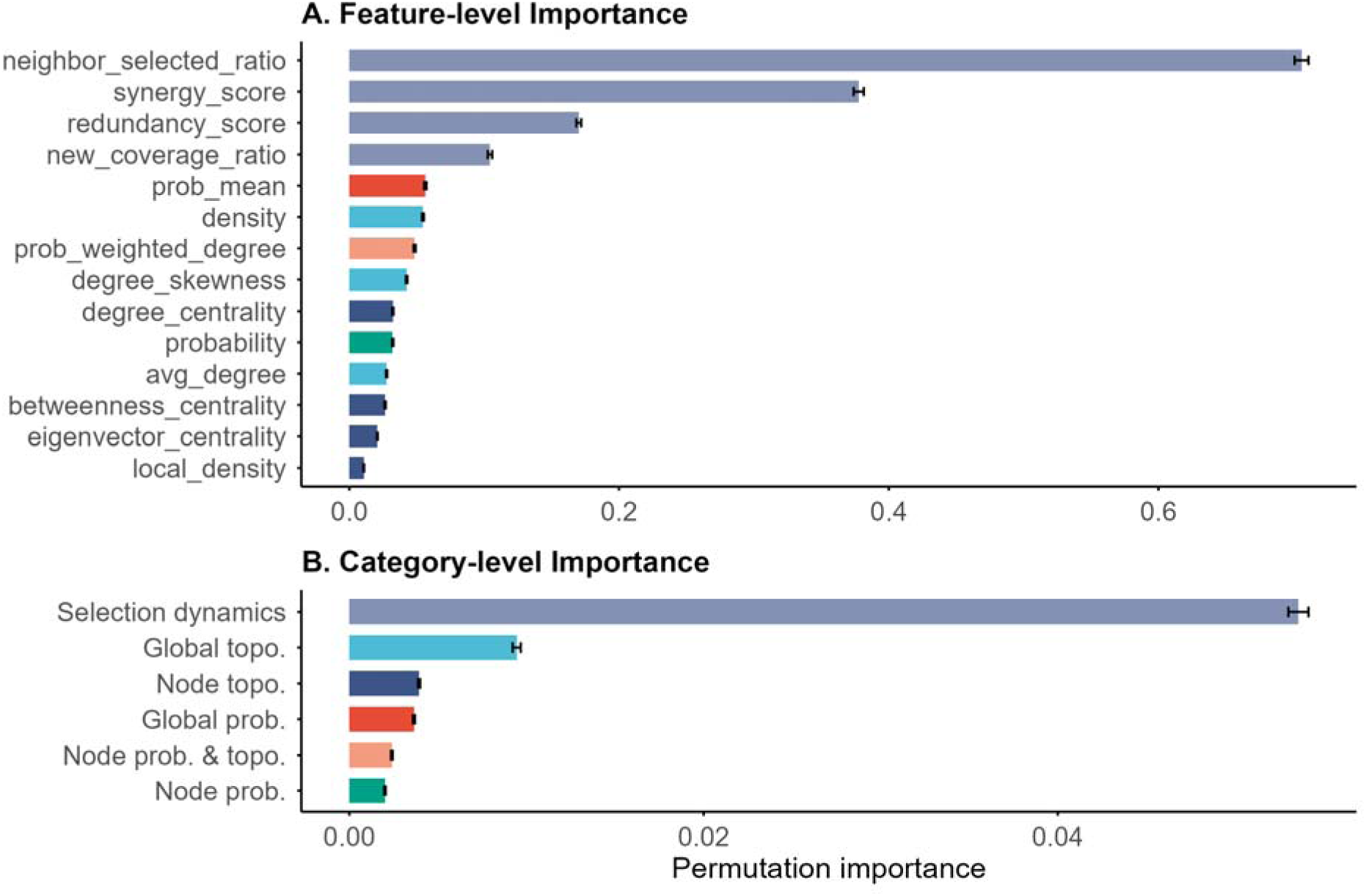
Overall feature and category importance of the RFSM. (A) Permutation importance of individual features, colored by category. (B) Grouped permutation importance of feature categories ordered by importance scores. Bars represent the mean permutation importance scores across 100 repetitions, with error bars showing the standard deviations.

#### Rank-specific feature and category importance

Stratifying feature importance by rank of selection revealed a clear shift in the determinants of early detection performance across selection stages (Fig. 9). For the first sentinel site, node and global-level network topology features dominated, with *betweenness_centrality* as the most important individual feature (Fig. S8). At later stages, dynamic selection features became the primary determinants, though their relative contribution gradually decreased with increasing selection rank (Figs. 9 and S8). Within this category, *neighbor_selected_ratio*, *redundancy_score* and *synergy_score* were more important than *new_coverage_ratio*. Node-level and global topology contributed modestly to early detection performance, but tended to gain slightly higher relative importance as additional sites were selected. In contrast, all emergence probability-related categories, including global and node emergence probability related, and node emergence probability-weighted topology measures, played only minor roles across all selection stages.

**Figure 9.**
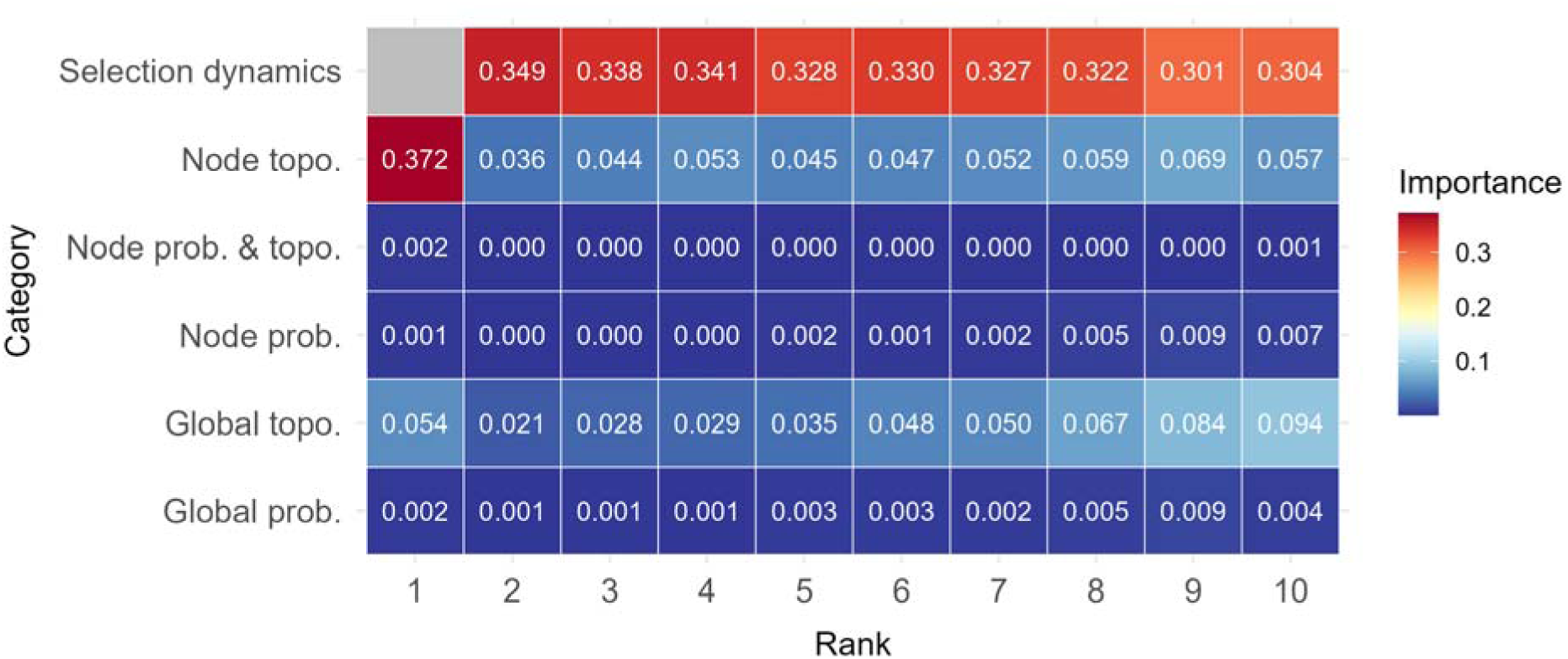
Rank-specific feature importance at the category level. Heatmap shows the contribution of each feature category across sentinel selection ranks. Rank-specific feature importance at the feature level can be found in Fig. S8.

#### Sensitivity analyses

Across all sensitivity analyses, selection dynamics, global network topology, and node topology consistently showed the highest overall importance, while emergence probability-related categories remained comparatively less influential (Fig. 10). However, their exact relative contributions varied across stratifications. When stratified by the total number of nodes (*N*, Fig. 10A) or the within-module connection probability (*p*, Fig. 10D), node topology became slightly more important, whereas selection dynamics and node emergence probability showed a modest decline with increasing values of *N* or *p*. In contrast, increasing mean degree (μ, Fig. 10B) led to a continuous increase in the contribution of selection dynamics, accompanied by a reduction in probability-related importance. Although emergence probability-related categories were generally less influential, their importance tended to rise with increasing degree heterogeneity (σ, Fig. 10C) and kurtosis of the emergence probability distribution (Fig. 10E), while the importance of selection dynamics first increased and then decreased. When stratified by the correlation between node degree and emergence probability (ρ), node emergence probability became less important as ρ increases (Fig. 10F). After stratification based on the basic reproduction number (*R*_0_), no differences were observed among categories (Fig. 10G). Individual feature-level results for each category are presented in Fig. S9.

**Figure 10.**
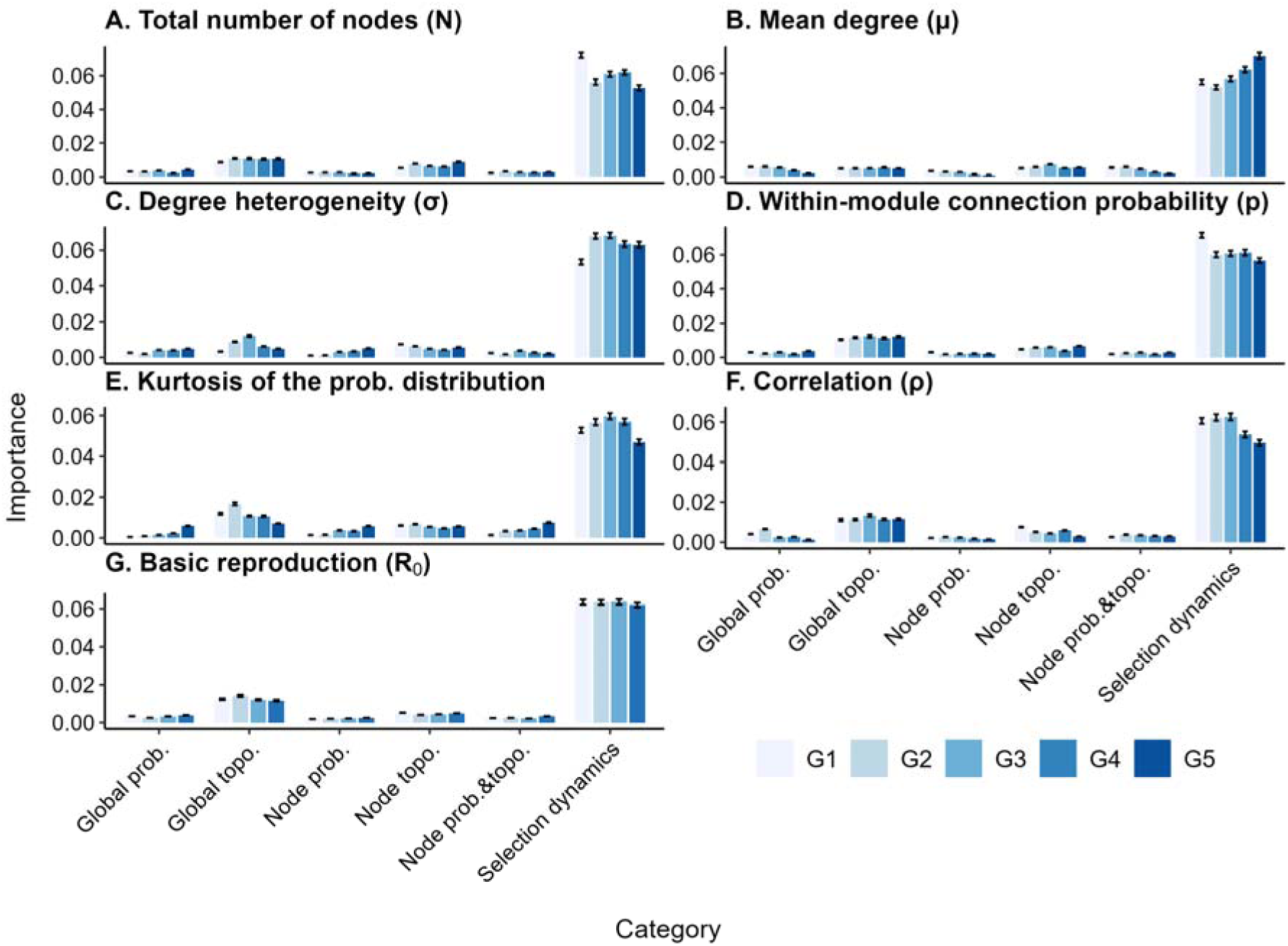
Sensitivity of category-level feature importance under different stratification parameters. Mean normalized importance of 6 categories across stratifications by (A) total number of nodes (*N*), (B) mean degree (μ), (C) degree heterogeneity (σ), (D) within-module connection probability (*p*), (E) kurtosis of the emergence probability distribution, (F) correlation between node degree and emergence probability (ρ), and (G) basic reproduction number (*R*_0_). Each bar represents the mean importance within one parameter group (G1–G5), with error bars indicating standard errors across 100 repetitions.

When stratified by rank, at rank=1, structural features were consistently the most important category across all groups (Fig. S10). From rank=2 onward, although the importance of selection dynamics showed a declining trend, it remains the most significant category, whereas the importance of emergence probability-related categories tended to rise. The relative importance among all categories varied with stratification parameters in a manner consistent with non-rank specific comparisons. However, when stratified by degree heterogeneity (σ) and kurtosis of emergence probability distribution, the trend of reduced node topological importance becomes more obvious than in the non-rank analysis (Fig. S10). Rank-specific feature importance sensitivity analysis stratified by parameters were shown in Figs. S11–S17.

Finally, when using different proportion of top-ranked nodes for model training, dynamic features remained the most influential. As the proportion increased, node topology became less while emergence probability became more important, possibly because lower-ranked nodes are noisier and less informative, since their objective function values are very close to each other (Figs. S18–19).

## Discussion

Nodes in complex networks often exhibit varying probabilities of being the initial seed for infectious disease outbreaks, determined by their environmental, behavioral or biological characteristics (e.g., animal markets, transportation hubs). This important heterogeneity is however typically overlooked in studies about the selection of sentinel surveillance nodes on complex networks. In this study, through simulations of outbreaks on both synthetic and empirical networks, we demonstrated that early detection performance was affected by both network topology and emergence probability, with the former playing a dominant role. We developed a computationally efficient random forest-based surrogate model for selecting sentinel nodes, validated its surveillance performance and computational feasibility across different networks, and implemented an online platform to facilitate its real-world applications. These findings highlight the moderate added value of incorporating node-specific emergence probabilities into sentinel surveillance design on complex networks and offer practical guidance for improving early outbreak detection.

Our findings confirmed the critical role of network topology in determining early detection performance. For example, topological and dynamic selection features consistently contributed more to node selection than features related solely to node or global emergence probability across all ranks (Figs. 8–10). Importantly, rank□specific analyses revealed a strategic shift: at the first selection stage, sentinel choice is dominated by topological features, favoring structurally central nodes such as transportation hubs or dense marketplaces, which align epidemiologically with “superspreader” roles [65]. As the sentinel set expands, however, dynamic selection features like neighbor□selected ratio and synergy score become increasingly decisive, identifying nodes that maximize coverage while mitigating overlap. For instance, neighbor□selected ratio quantifies neighboring nodes already selected, thereby guiding an efficient, non□redundant expansion of surveillance coverage. This shift mirrors adaptive, subregional surveillance strategies and resonates with modeling studies advocating dynamic intervention on networks [66–68].

Although emergence probability-related features were consistently less influential, they still provided meaningful contributions, particularly under conditions of high degree heterogeneity or uneven emergence probability distributions (Fig. 10). These features capture spatial or nodal variability in outbreak likelihoods, which can refine sentinel placement by highlighting regions with elevated emergence potential. However, their overall importance remained lower than topological or dynamic features because network connectivity largely governs transmission reach and detection opportunities, often overshadowing intrinsic emergence probabilities. Moreover, when emergence probability strongly correlates with node degree, its information content becomes partially redundant with structural attributes. Thus, effective surveillance design ultimately depends more on structural integration and dynamic adaptability than on static emergence probability.

The strong predictive performance of the RFSM indicates that data-driven surrogate modeling can effectively approximate the behavior of optimization-based approaches in sentinel site selection. By integrating key structural and dynamic features, the RFSM achieves near-optimal early detection performance comparable to the greedy algorithm while maintaining robust generalization across diverse networks. Its accuracy is particularly reliable for the early stages of sentinel selection, where it was trained to emulate the greedy algorithm’s decisions, but its predictions become less precise in later selection steps as the underlying relationships between features and performance weaken. As with most data-driven models, the RFSM’s accuracy may also decline when applied to networks whose structural or epidemiological relationships deviate substantially from those represented in the training data. Furthermore, the effectiveness of different sentinel selection strategies, including RFSM, was also closely tied to the heterogeneity of network structure. For instance, in the case of the university network, where high-degree nodes are closely interconnected, the *global* strategy performed even worse than the *Random* strategy as the number of sentinel nodes increased, likely due to the selection of neighboring nodes that capture essentially redundant information (Fig. S4C). This observation highlights the importance of understanding network structure when designing surveillance strategies, as densely interconnected high-degree nodes may lead to information overlap, reducing the efficiency gains of topology-based strategies in heterogeneous networks [69,70]. Nevertheless, the RFSM provides a practical balance between accuracy and computational efficiency, making it particularly valuable for scalable or time-sensitive surveillance design.

Although the *greedy*, *GA*, and RFSM strategies all achieved high performance in early outbreak detection, their computational burden differed substantially. The *greedy* algorithm iteratively evaluated all remaining candidate nodes (*n*) at each of the *l* selection steps through *s* Monte Carlo simulations (Eq(1)), resulting in a time complexity of O(*n*×*l*×*s*). This process quickly becomes infeasible as network size or the number of simulations increases. GA further amplifies this computational demand by evaluating multiple candidate solutions per generation and often requires hundreds of generations to converge. In contrast, the RFSM replaces these iterative simulations with predictive inference using a pre-trained model that performs well even on unseen networks. Each selection step only requires computing node-level features and performing predictions, yielding a linear complexity of O(*n*×*l*). This substantial reduction in computational cost highlights RFSM’s suitability for large-scale, dynamic, or real-time surveillance applications where traditional optimization methods are computationally prohibitive.

A key challenge in network epidemiology is the difficulty of obtaining the complete contact networks. Our analysis showed that sentinels selected from highly incomplete networks (e.g., missing 50% of edges) by RFSM performed nearly as well as those selected from complete networks, which implies that disease surveillance systems can be effectively designed even using data from rapid, partial assessments or sampled surveys, without requiring a full mapping of the contact network, thereby lowering barriers to implementation. This robustness likely arises because RFSM captures robust topological roles (such as bridge nodes or local connectivity patterns) rather than overfitting to specific, fragile connections. In contrast, strategies that rely solely on identifying the highest-degree nodes (*global*) can be more brittle if the true hubs are unobserved.

Our sensitivity analyses demonstrate how network topology and emergence probability characteristics affect the contribution of different features (Figs. 10 and S9–17). We found that the importance of selection dynamics increased with higher mean degree (μ) (Figs. 10B and S9), likely because nodes in more highly connected networks tend to convey overlapping information, which should be minimized during the node selection process. The rising importance of emergence probability-related features under greater degree heterogeneity (σ) and higher kurtosis of emergence probability distribution indicates that when risks are highly uneven or heavy-tailed, node-level emergence variability becomes more informative for monitoring placement. However, when degree and emergence probability are strongly correlated (ρ), this information is largely redundant with structural metrics, leading to reduced relative importance. The absence of clear *R*_0_ effects suggests that variations in transmission potential do not fundamentally shift the structural-dynamic balance.

Our results also suggest that monitoring a small number of strategically selected nodes can achieve a high level of surveillance performance, as adding more sentinels yields progressively smaller performance gains. More specifically, we found that monitoring just six nodes can capture about 90% of the performance of the complete surveillance network monitoring all nodes, and notably, the required number of sentinel nodes did not scale with network size (Fig. 7). This conclusion is consistent with a global wastewater surveillance study, which found that 10–20 optimally placed wastewater sentinel sites across more than 4,000 international airports were sufficient to provide timely situational awareness [71]. One possible explanation is that, when sentinel nodes cover nodes with high emergence probabilities, even a small number of nodes can effectively interrupt the most potential transmission chains.

Several limitations must be acknowledged when interpretating the results. First, we assumed that node-specific emergence probabilities are known, which may not be directly observable in practice. Although some studies have attempted to infer spatial heterogeneity in emergence probabilities based on past emerging infectious disease events and factors like land use patterns, human population density, and wildlife and livestock distributions [1,6,11], more detailed data, such as wildlife habitat, villagers’ hunting trajectories, and forest contact frequencies, can be further incorporated to better characterize the spatial distribution of EID emergence probabilities [72,73]. Second, we represented contact patterns as unweighted, undirected and static graphs, which may not fully capture the complexity of real-world interactions. Extending the RFSM framework to temporal [29], weighted, or multilayer networks [74,75] would further enhance its applicability to practical surveillance system design. Third, we assumed that the cost of placing each sentinel node is the same. Future work can assign different costs to different nodes based on their accessibility and introduce constraints on total cost or incorporate total cost as an additional objective in a multi-objective optimization framework. Fourth, our simulations considered only single-seed outbreaks, whereas in reality multiple introductions may occur simultaneously or repeatedly. Under such scenarios, the relative importance of emergence probabilities may increase, as multiple seeding events amplify the influence of initial outbreak locations on surveillance performance. Finally, we focused exclusively on respiratory infections and used the simplest SIR model, assuming perfect detection for the infectious nodes. Future studies can incorporate more complex transmission dynamics and detection rate to improve the generalizability of the framework to a wider range of infectious diseases.

In conclusion, through simulations and machine learning models, we found that node-specific emergence probabilities significantly enhance early outbreak detection performance, though their influence is secondary to network topology. Notably, approximately 90 percent of the surveillance performance can be achieved with as few as six sentinel nodes, and the required sentinel size does not scale with network size. Leveraging key node characteristics identified in our analysis, we developed a computationally efficient surrogate model for optimal sentinel selection on complex networks and validated its surveillance and computational performance on empirical networks. This model was further implemented as an online platform. Our findings underscore the importance of accounting for heterogeneity in node-specific emergence probability when designing sentinel surveillance system on complex networks and provide a practical and scalable solution that can be readily applied in real-world settings, especially when resources and technical capacity are limited.

## Data Availability

All data produced in the present study are available upon reasonable request to the authors

