## Supplemental Figures for "Network topology outweighs emergence probability in surveillance sentinel placement"

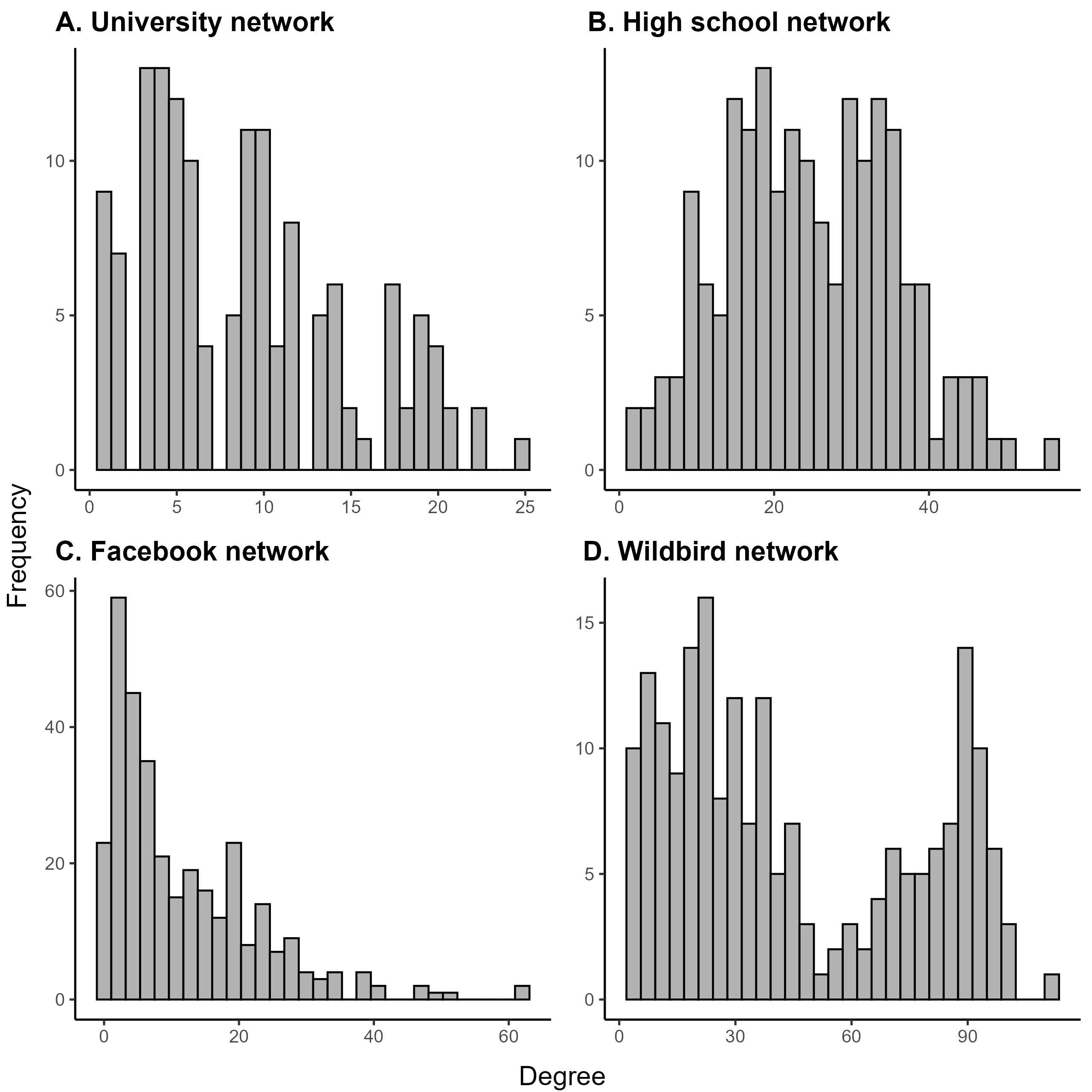
Figure.S1

**Figure S1. Degree distributions of empirical networks used in this study.** Histograms showing the frequency distribution of node degrees across four real-world networks: (A) University network, (B) High school network, (C) Facebook network, and (D) Wild bird network.

Figure.S2

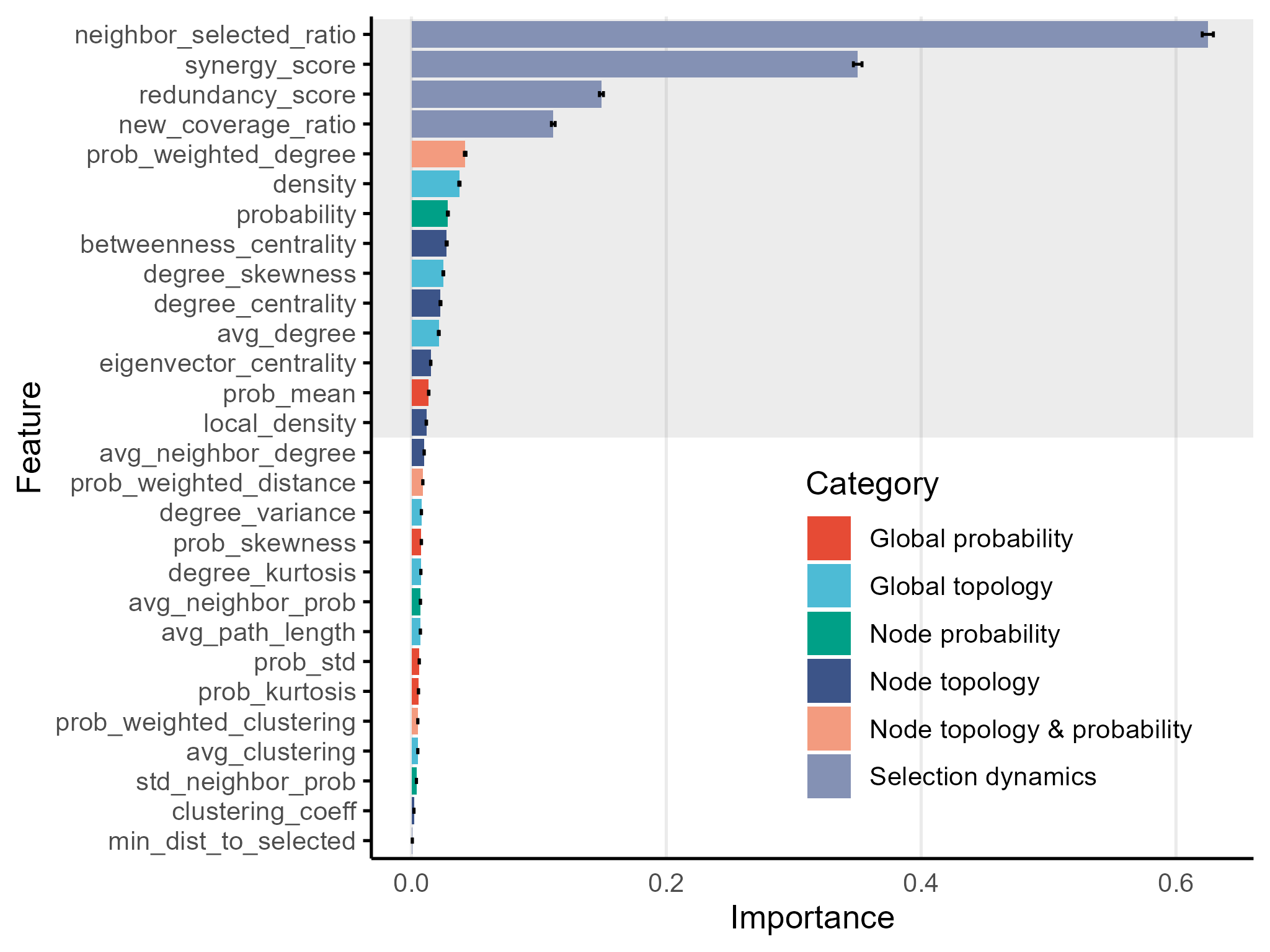

**Figure S2. Feature selection of the RFSM.** Permutation importance of all candidate features, colored by category. Bars represent the mean importance scores across 100 cross-validations, with error bars showing the standard deviations. The shaded area highlights the final 14 features selected for model fitting.

Figure.S3

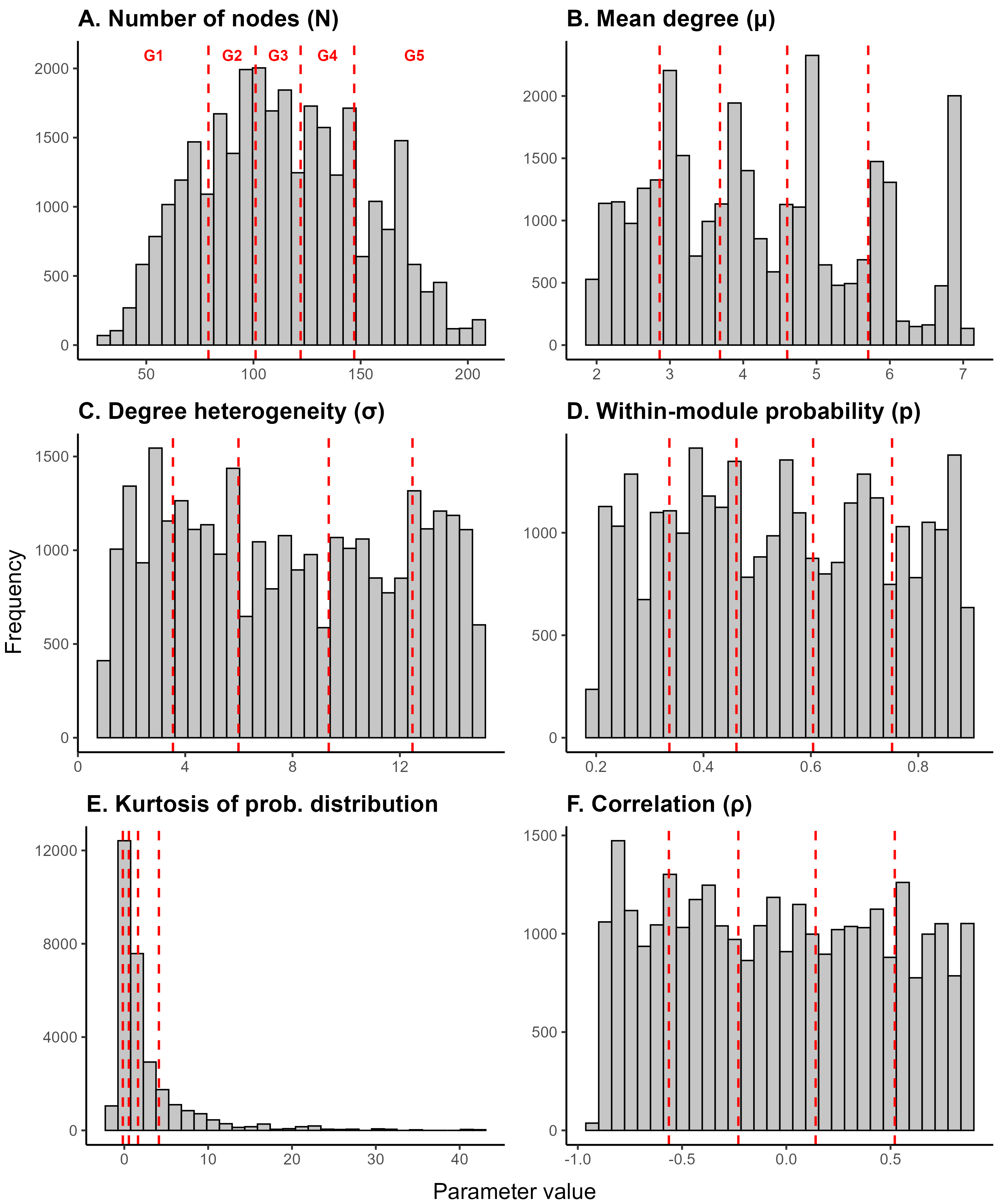

**Figure S3. The distribution of six parameters with quintile-based grouping.** Red dashed lines indicate quintile boundaries dividing each parameter into five groups (G1-G5).

Figure.S4

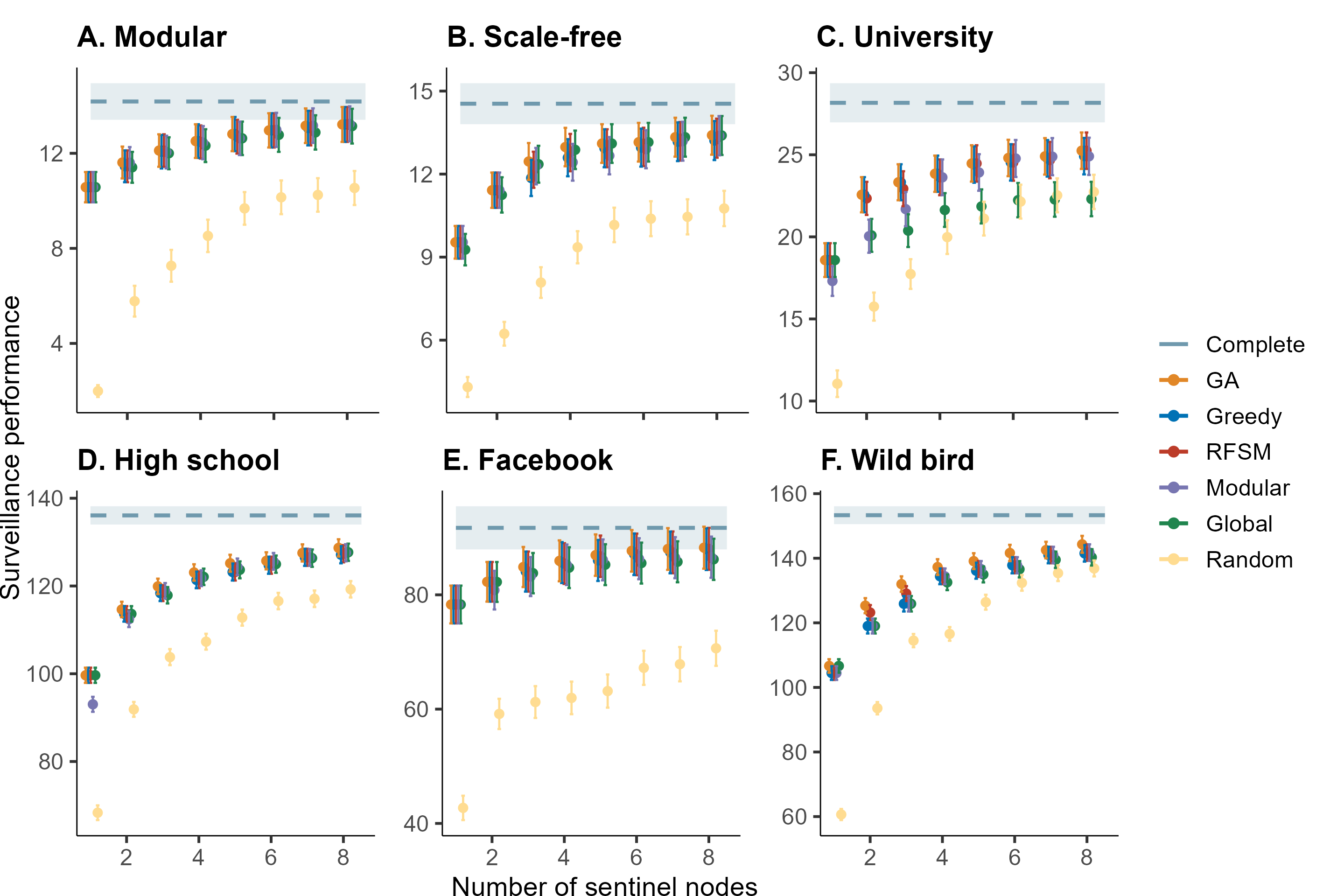

**Figure S4. Surveillance performance comparison across six networks with varying topological characteristics.** Each panel shows the surveillance performance (y-axis) as a function of the number of sentinel nodes (x-axis) for different node selection strategies. The blue dashed line represents the surveillance performance of monitoring all node in networks. Error bars indicate standard deviation across 100 simulation runs. For each network, node emergence probabilities were sampled from Beta(0.1, 5) distribution, with correlation coefficient (*ρ*) of -0.7 between node degree and emergence probability, and basic reproduction number (*R*_0_) set to 3.0.

Figure.S5

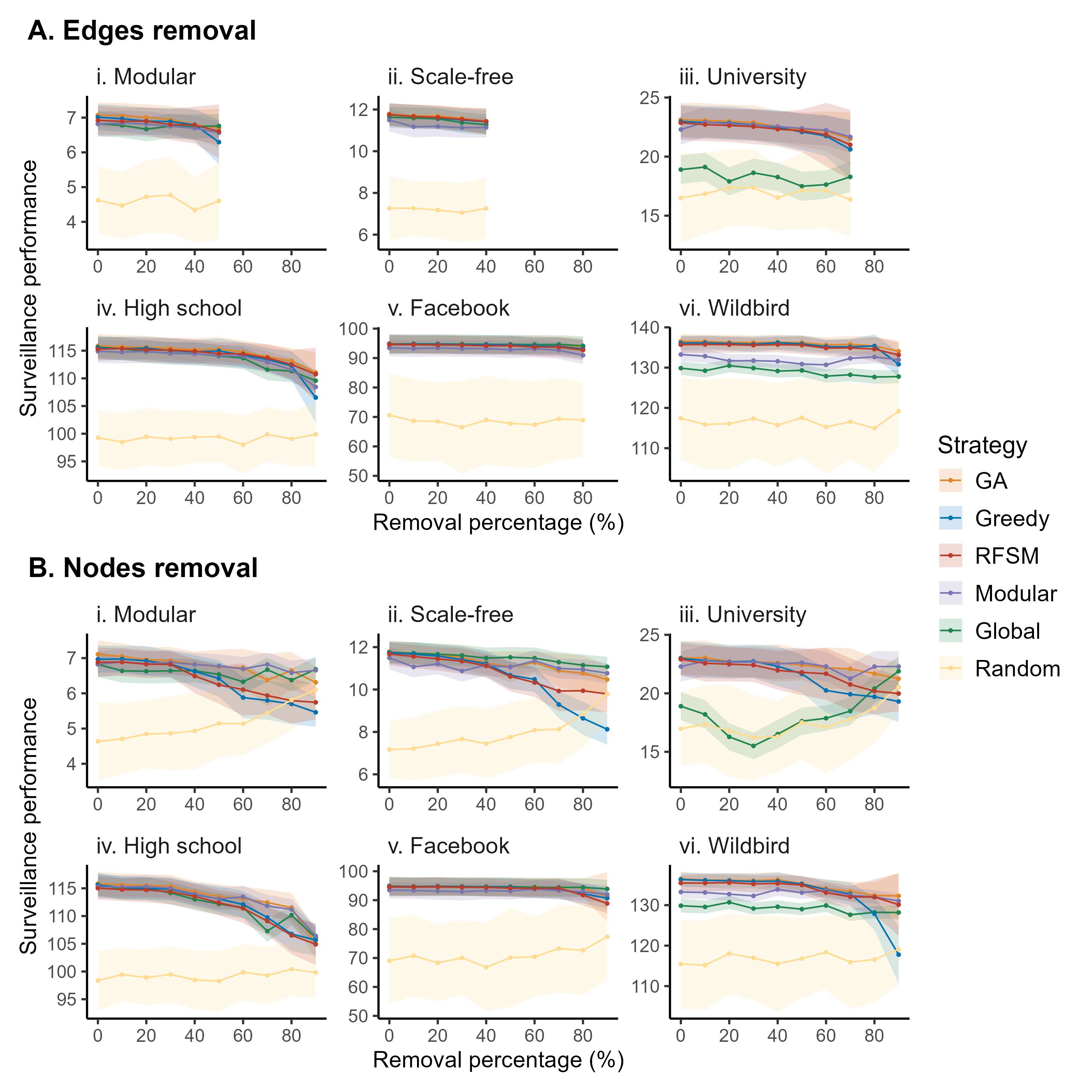

**Figure S5. Surveillance performance under incomplete network structure observation when 3 sentinel nodes were selected.** (A) shows results for edge removal, while (B) shows results for node removal. Columns (i–vi) correspond to different network types: Modular, Scale-free, University, High school, Facebook, and Wildbird. For each panel, the x-axis denotes the proportion of removed edges or nodes, and the y-axis represents mean surveillance performance. Solid lines indicate average performance across simulations, and shaded areas represent variability across10 repetitions.

Figure.S6

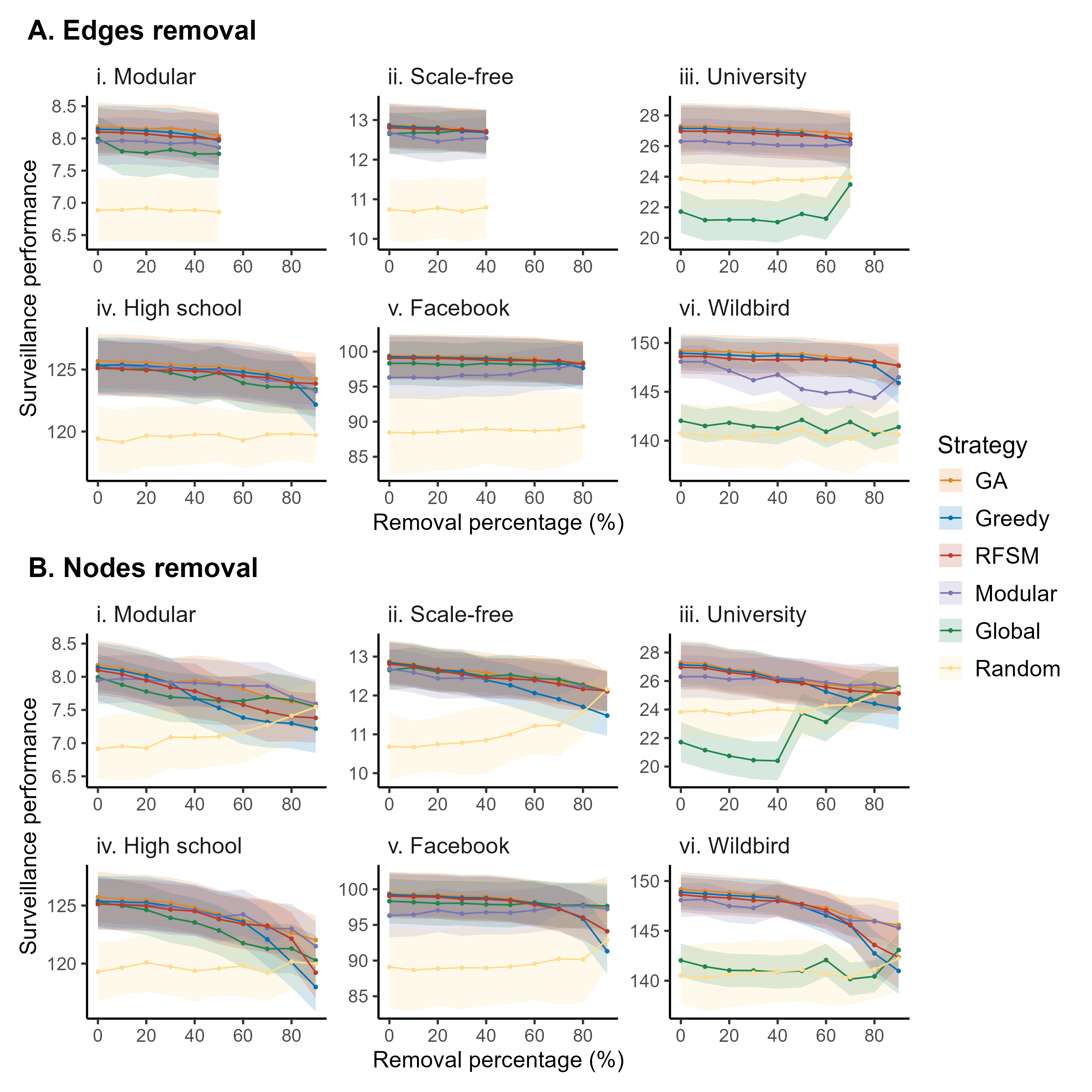

**Figure S6.** **Surveillance performance under incomplete network structure observation when 9 sentinel nodes were selected.** (A) shows results for edge removal, while (B) shows results for node removal. Columns (i–vi) correspond to different network types: Modular, Scale-free, University, High school, Facebook, and Wildbird. For each panel, the x-axis denotes the proportion of removed edges or nodes, and the y-axis represents mean surveillance performance. Solid lines indicate average performance across simulations, and shaded areas represent variability across10 repetitions.

Figure.S7

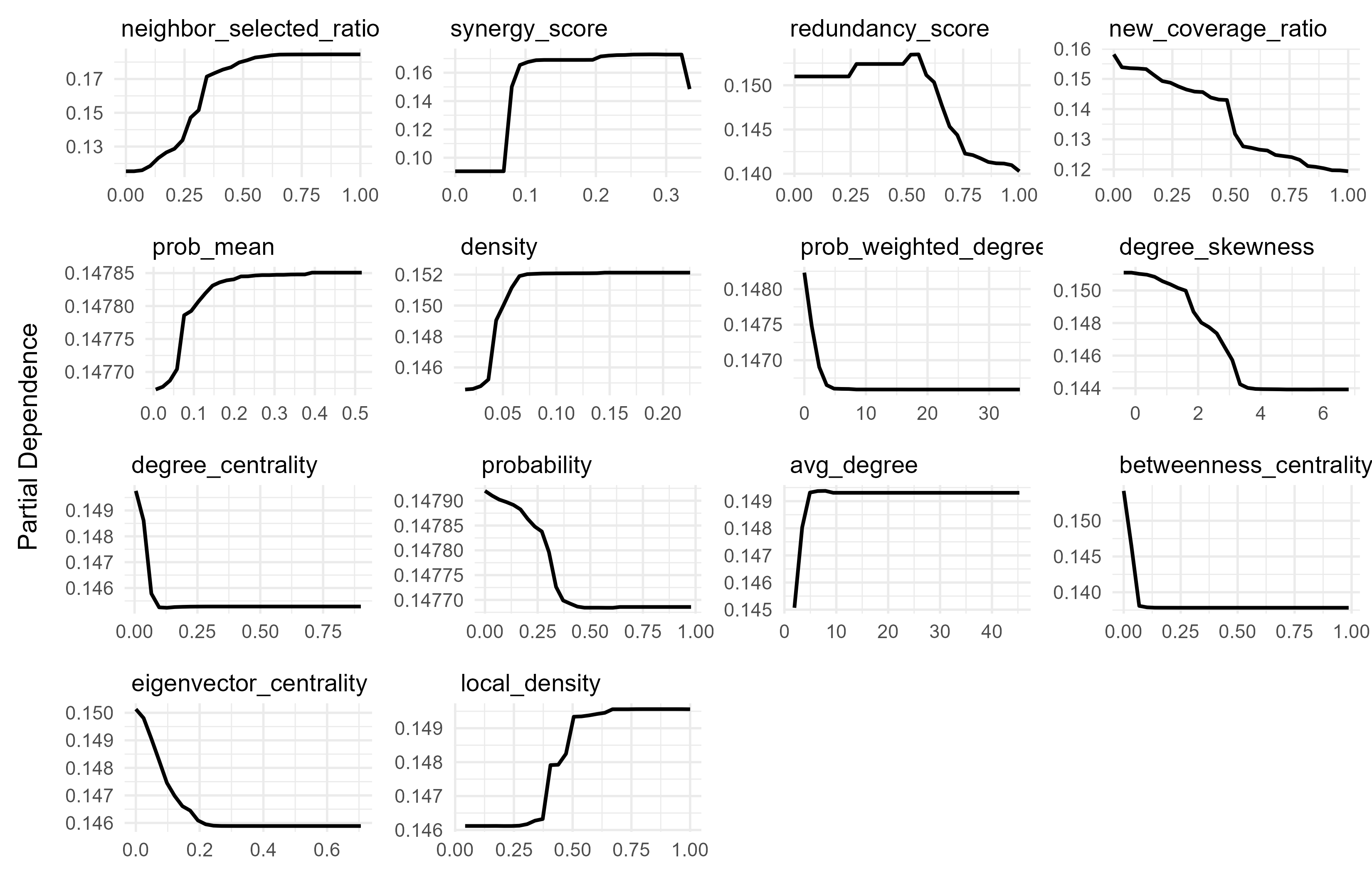

**Figure S7. Partial dependence plots of model-predicted relative rank for candidate sentinel nodes.** The model output is relative rank, with smaller values indicating better sentinel node selections.

Figure.S8

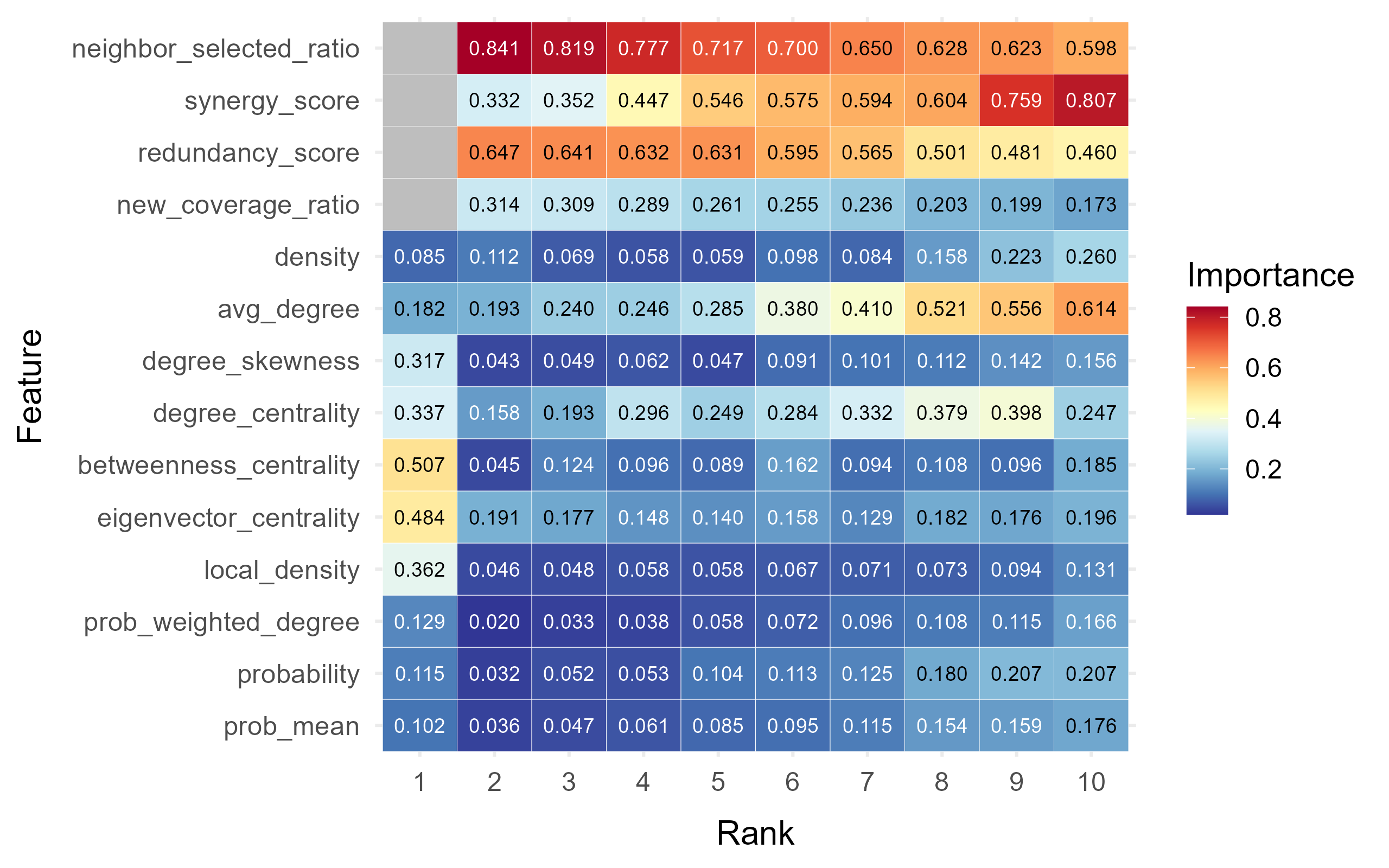

**Figure S8. Rank-specific feature importance at the feature level.** Heatmap shows the contribution of each feature across sentinel selection ranks.

Figure.S9

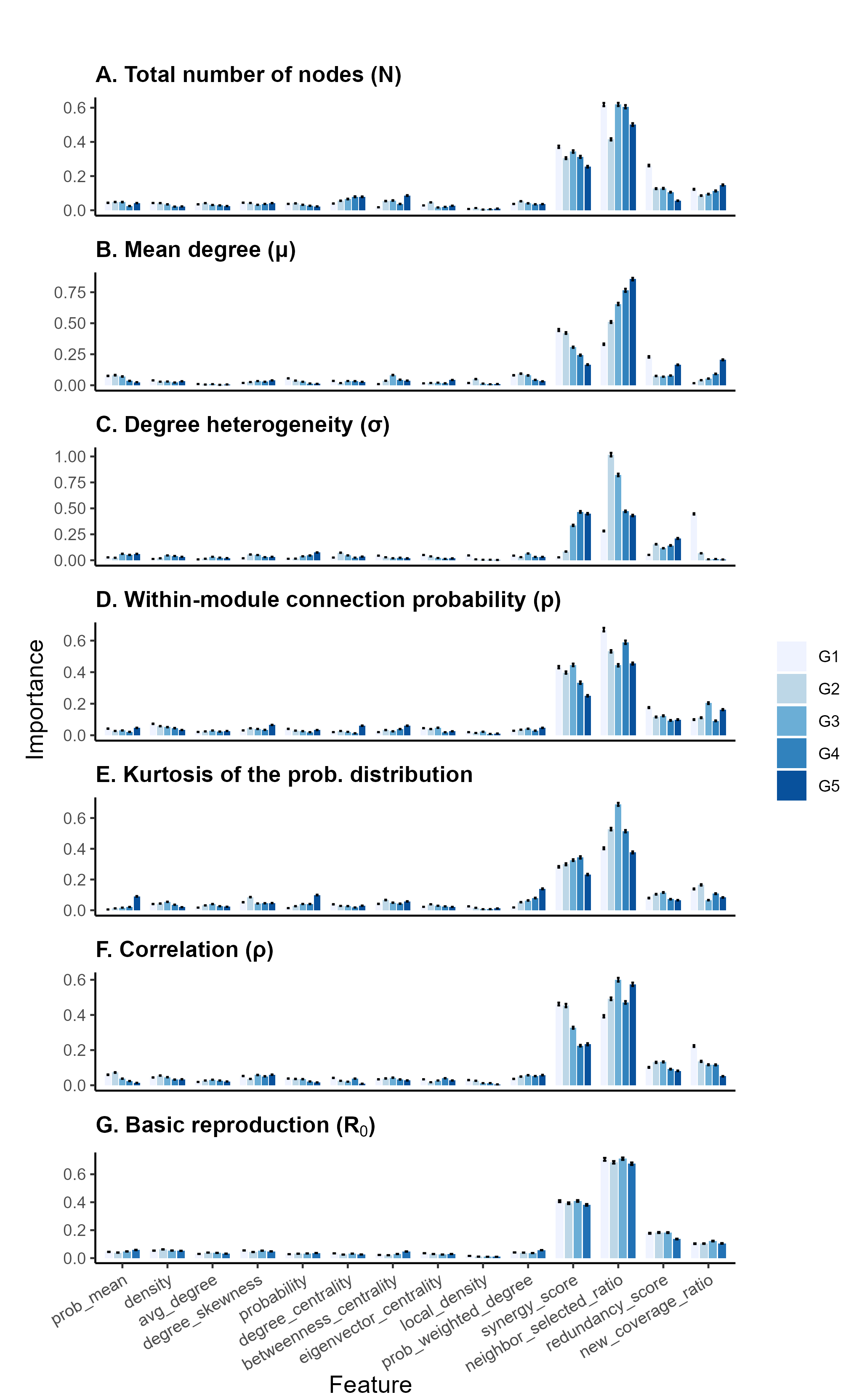

**Figure S9. Sensitivity of feature-level feature importance under different stratification parameters.** Mean normalized importance of 14 features across stratifications by (A) total number of nodes (*N*), (B) mean degree (*μ*), (C) degree heterogeneity (*σ*), (D) within-module connection probability (*p*), (E) kurtosis of the emergence probability distribution, (F) correlation between node degree and emergence probability (*ρ*), and (G) basic reproduction number (*R*_0_). Each bar represents the mean importance within one parameter group (G1–G5), with error bars indicating standard errors across 100 repetitions.

Figure.S10

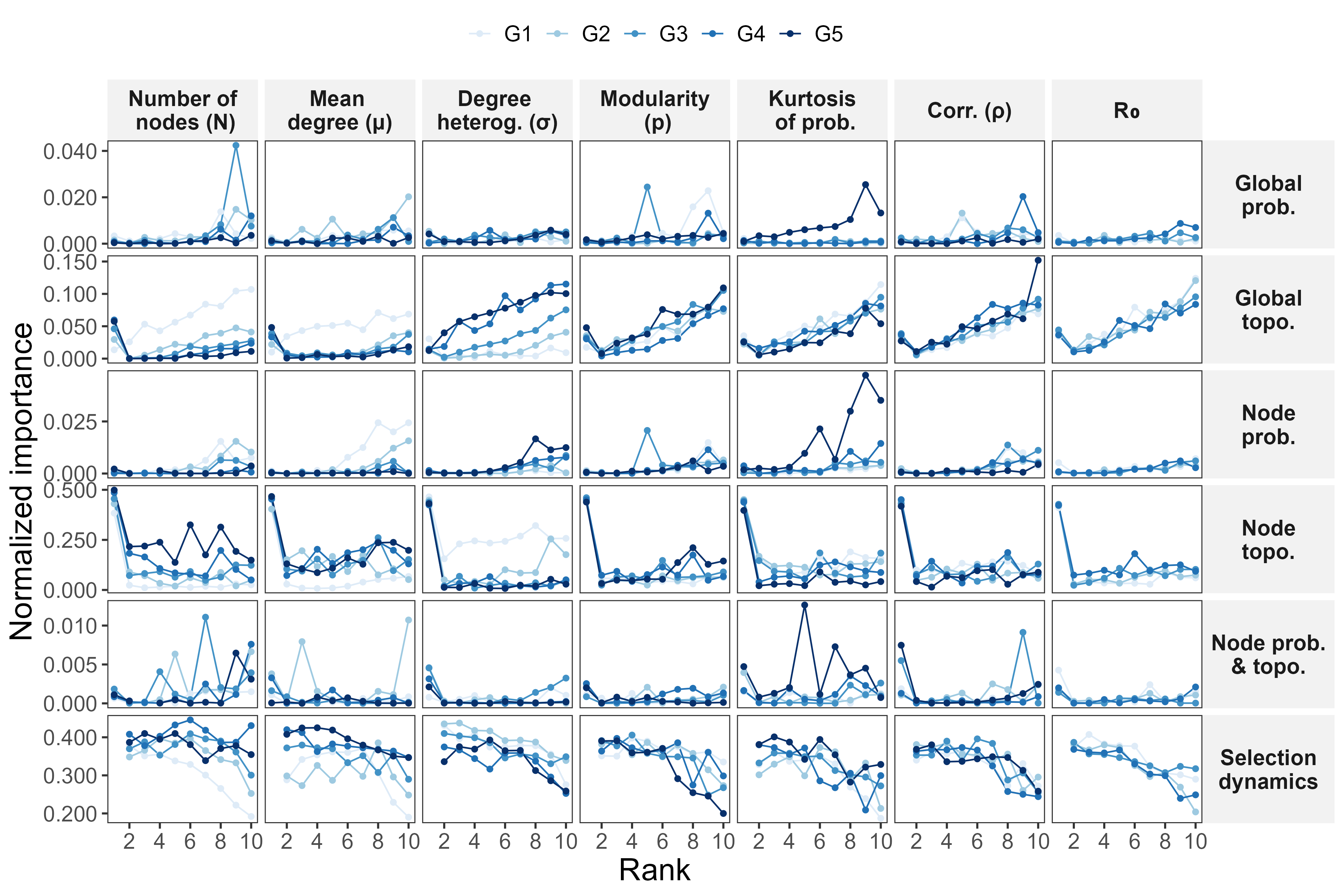

**Figure S10. Rank-specific sensitivity of category-level feature importance under different stratification parameters.** Normalized importance of 6 categories across ranks (x-axis) and stratifications (columns), including total number of nodes (*N*), mean degree (*μ*), degree heterogeneity (*σ*), within-module connection probability (*p*), kurtosis of the emergence probability distribution, correlation between node degree and emergence probability (*ρ*), and basic reproduction number (*R*_0_). Each row represents one feature category, and lines (G1–G5) denote stratified groups under each parameter.

Figure.S11

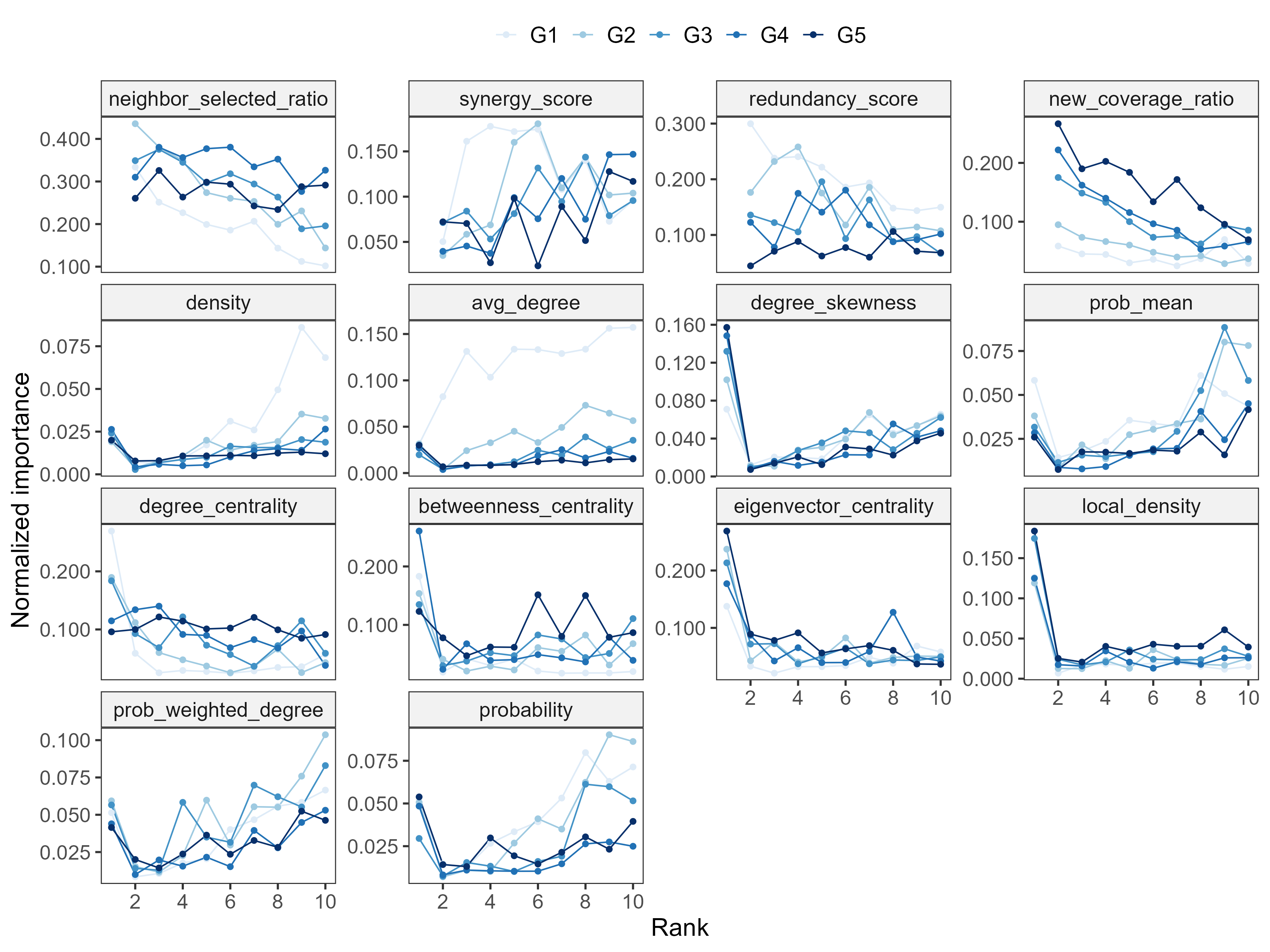

**Figure S11. Rank-specific sensitivity analysis of feature-level importance by total number of nodes (*N*).** Normalized feature importance across ranks (x-axis) under different network sizes (G1–G5). Each panel represents one feature included in the RFSM.

Figure.S12

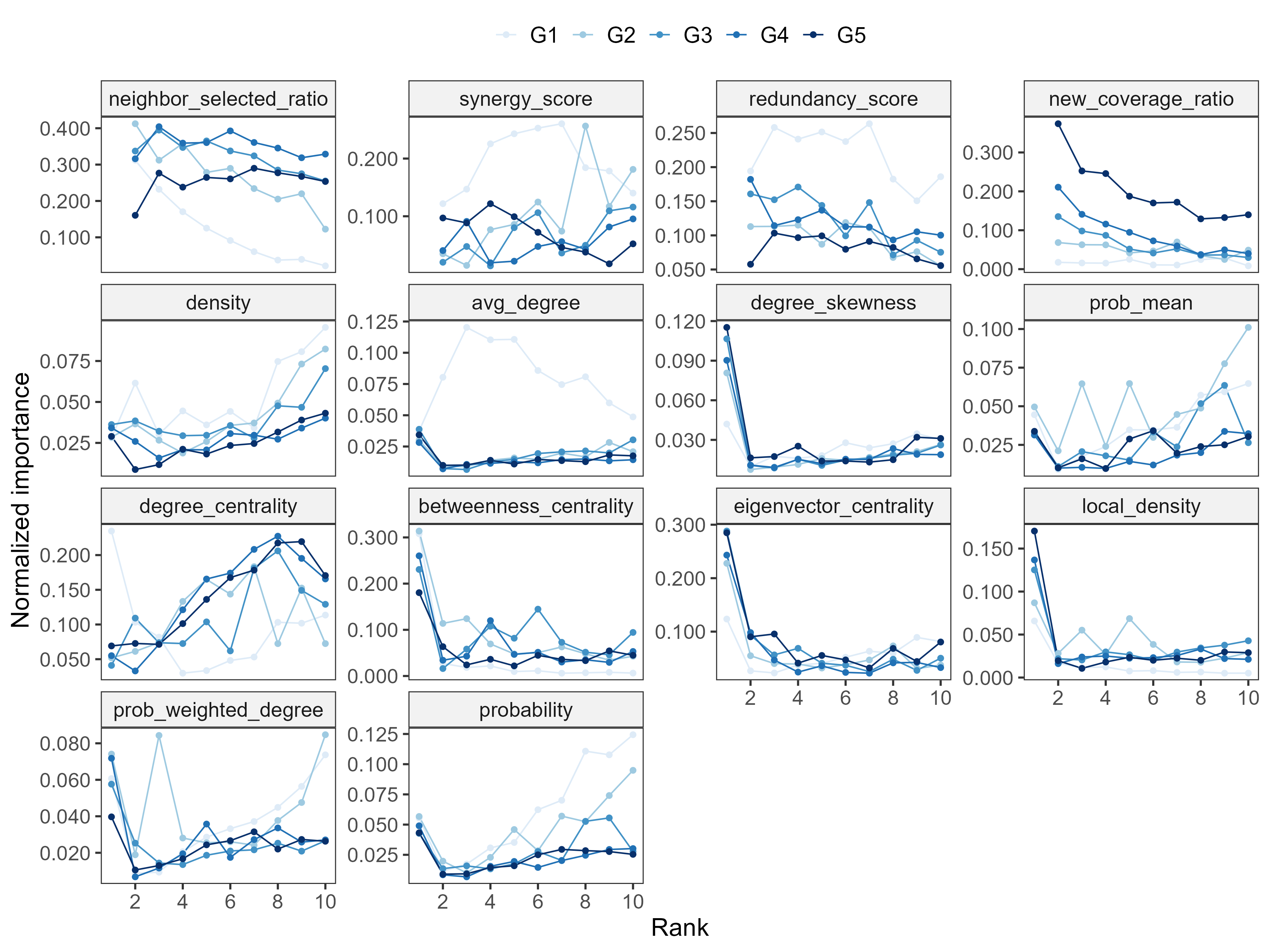

**Figure S12. Rank-specific sensitivity analysis of feature-level importance by mean degree (*μ*).** Normalized feature importance across ranks (x-axis) under varying mean degrees (G1–G5). Each panel represents one feature included in the RFSM.

Figure.S13

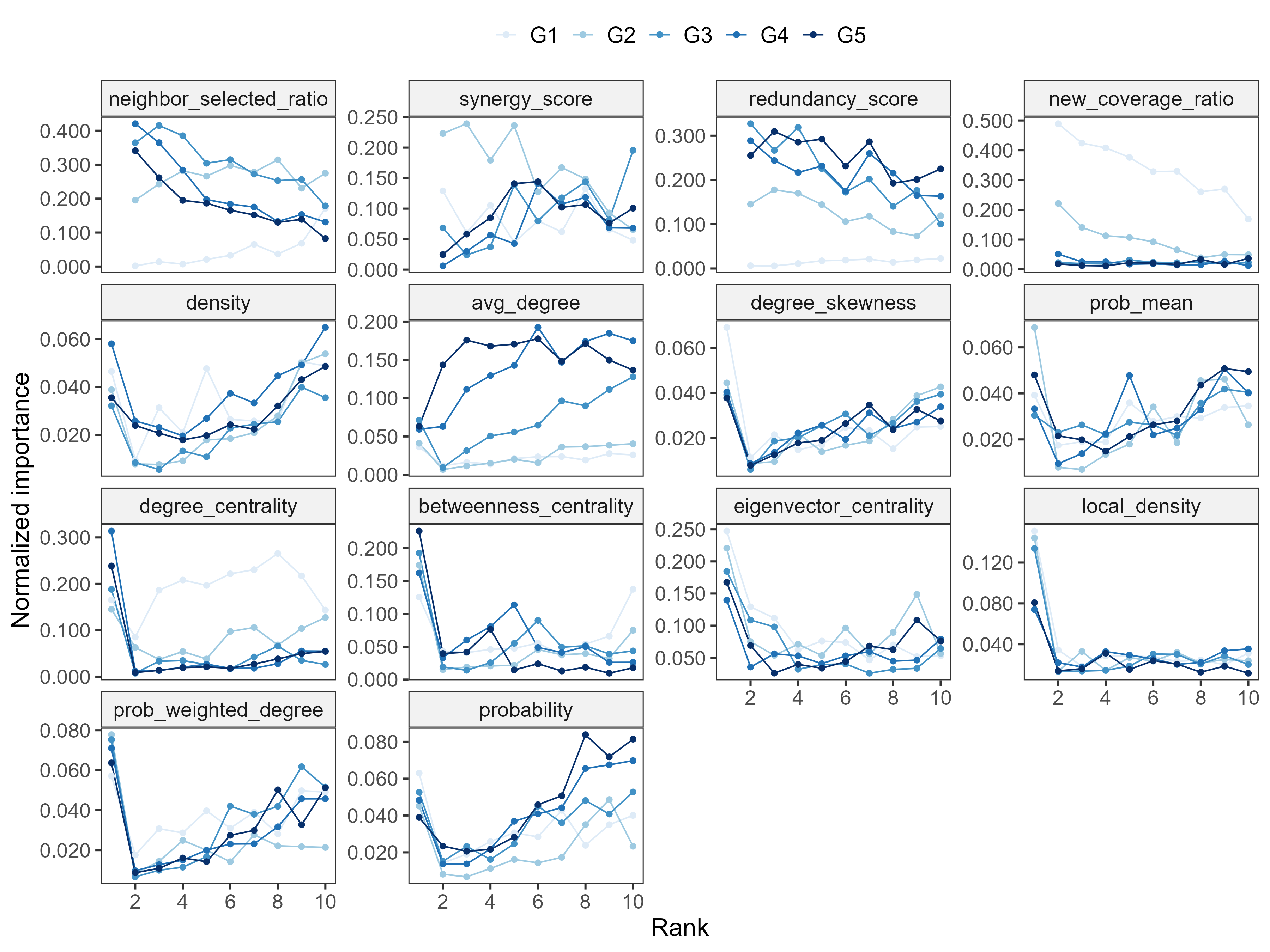

**Figure S13. Rank-specific sensitivity analysis of feature-level importance stratified by degree heterogeneity (*σ*).** Normalized feature importance across ranks (x-axis) under different degree heterogeneity (G1–G5). Each panel represents one feature included in the RFSM.

Figure.S14

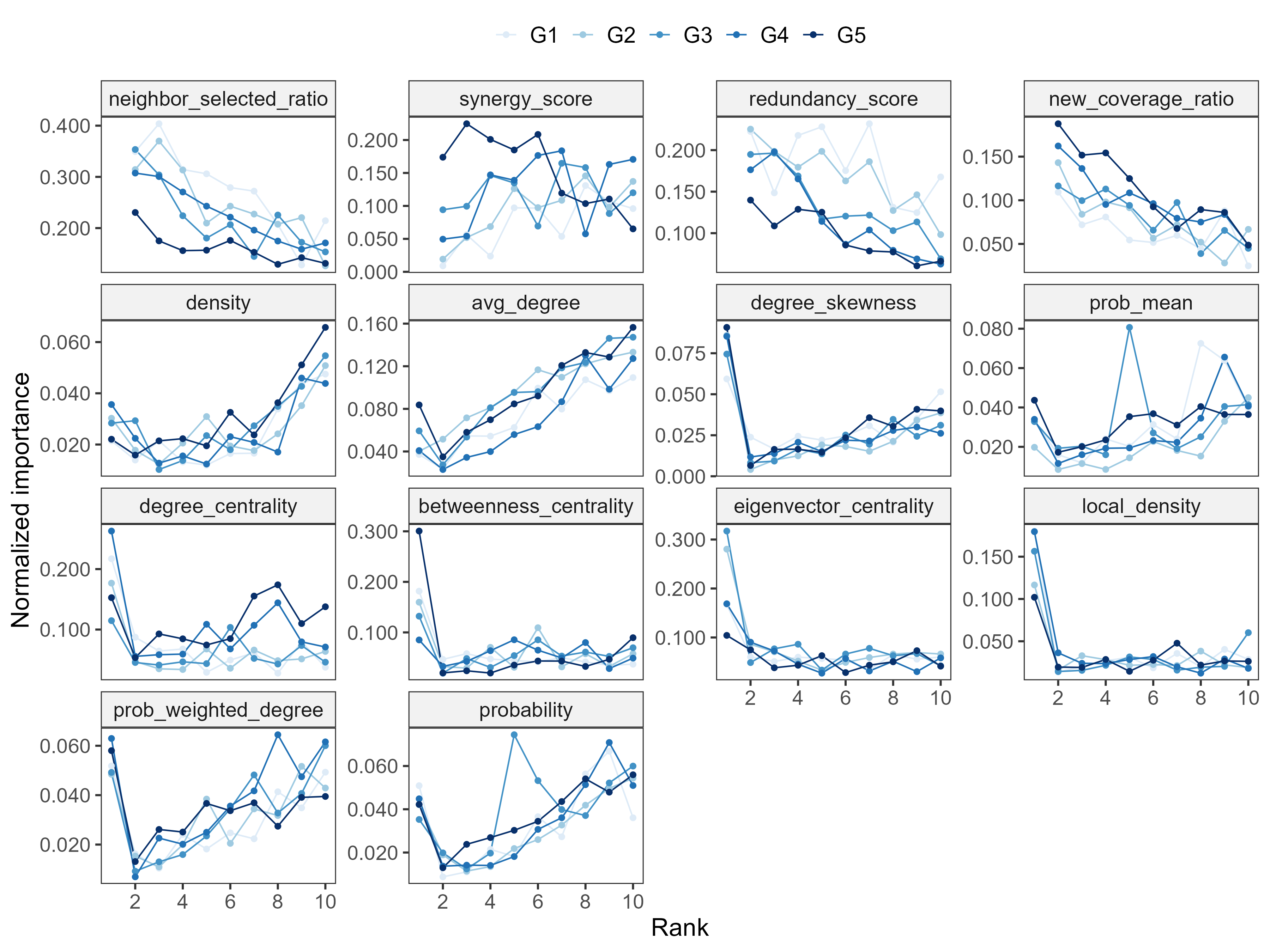

**Figure S14.** **Rank-specific sensitivity analysis of feature-level importance stratified by within-module connection probability (*p*).** Normalized feature importance across ranks (x-axis) under varying intra-module connectivity (G1–G5). Each panel represents one feature included in the RFSM.

Figure.S15

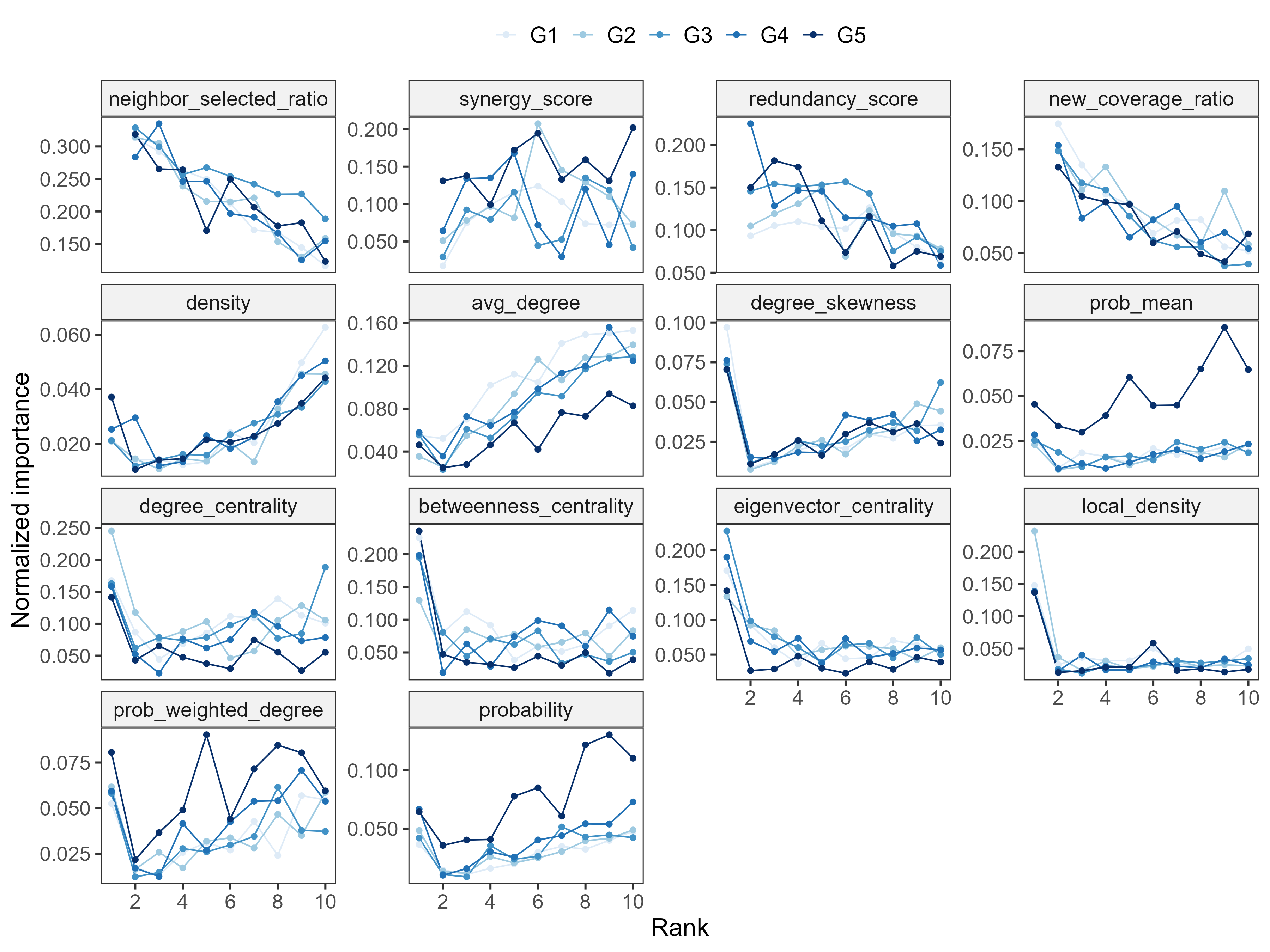

**Figure S15. Rank-specific sensitivity analysis of feature-level importance stratified by kurtosis of the emergence probability distribution.** Normalized feature importance across ranks (x-axis) under different kurtosis levels (G1–G5). Each panel represents one feature included in the RFSM.

Figure.S16

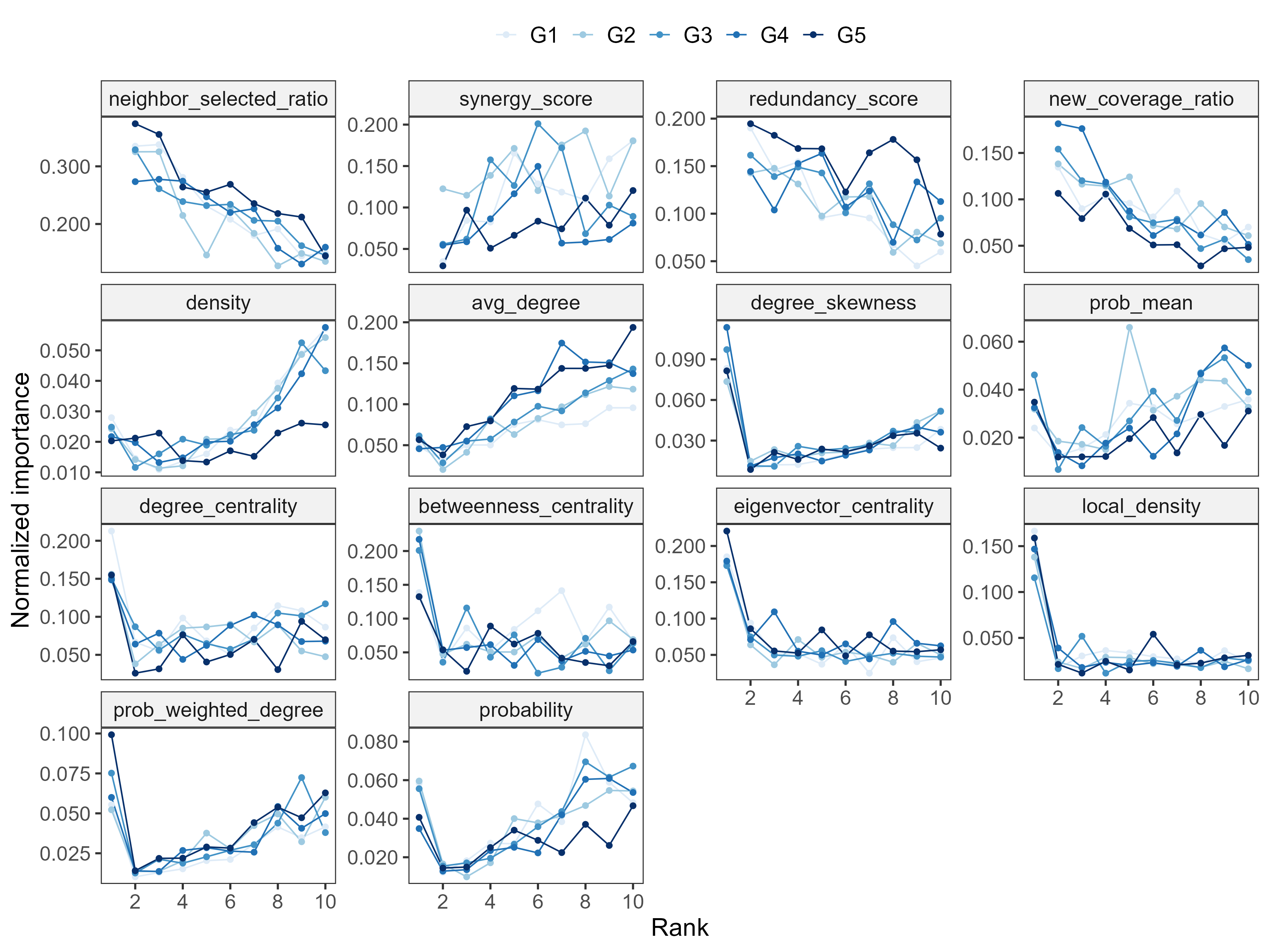

**Figure S16. Rank-specific sensitivity analysis of feature-level importance stratified by the correlation between node degree and emergence probability (*ρ*).** Normalized feature importance across ranks (x-axis) under varying correlation strengths (G1–G5). Each panel represents one feature included in the RFSM.

Figure.S17

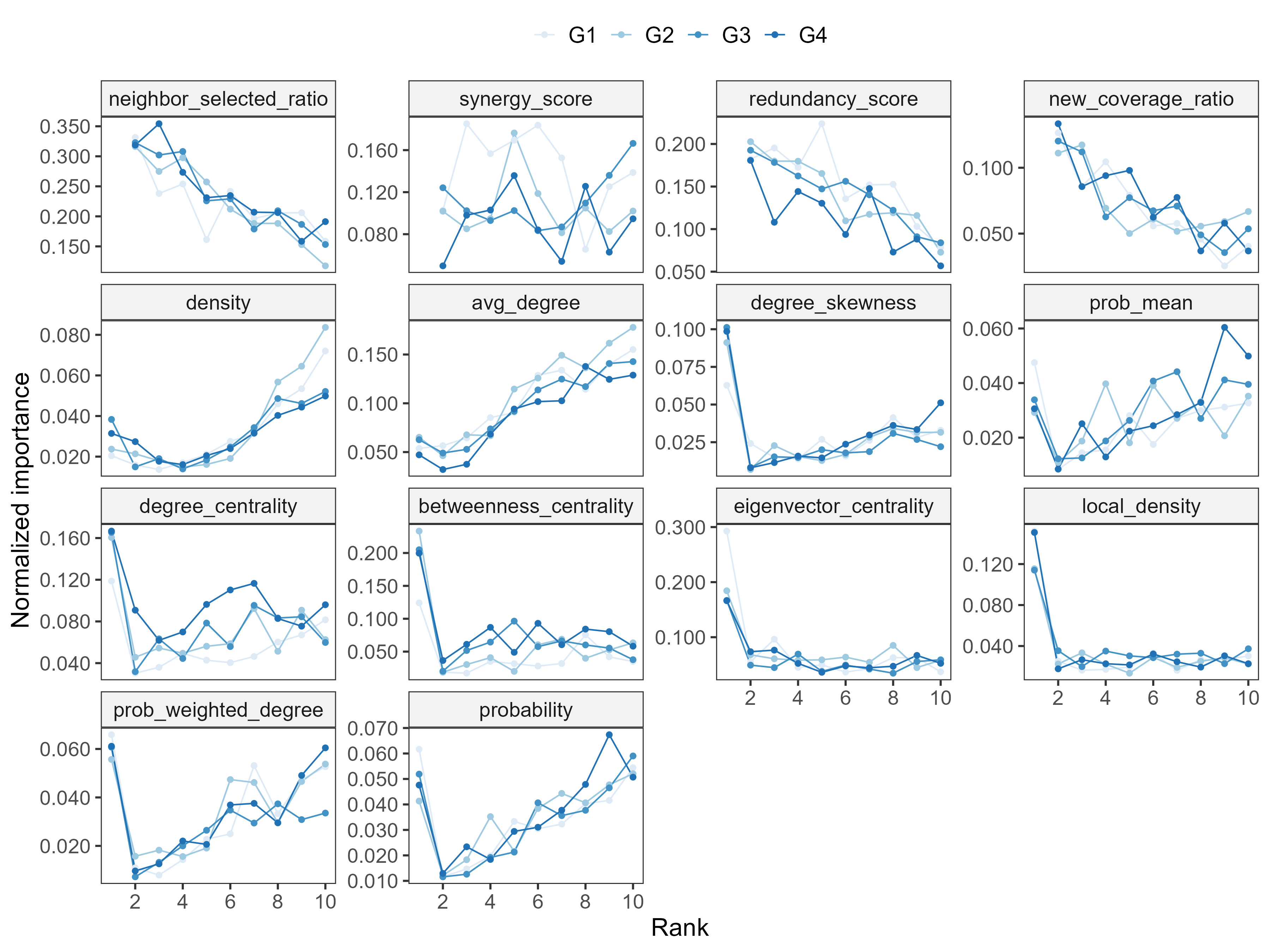

**Figure S17. Rank-specific sensitivity analysis of feature-level importance stratified by the basic reproduction (*R*_0_).** Normalized feature importance across ranks (x-axis) under different *R*_0_ values (G1–G4). Each panel represents one feature included in the RFSM.

Figure.S18

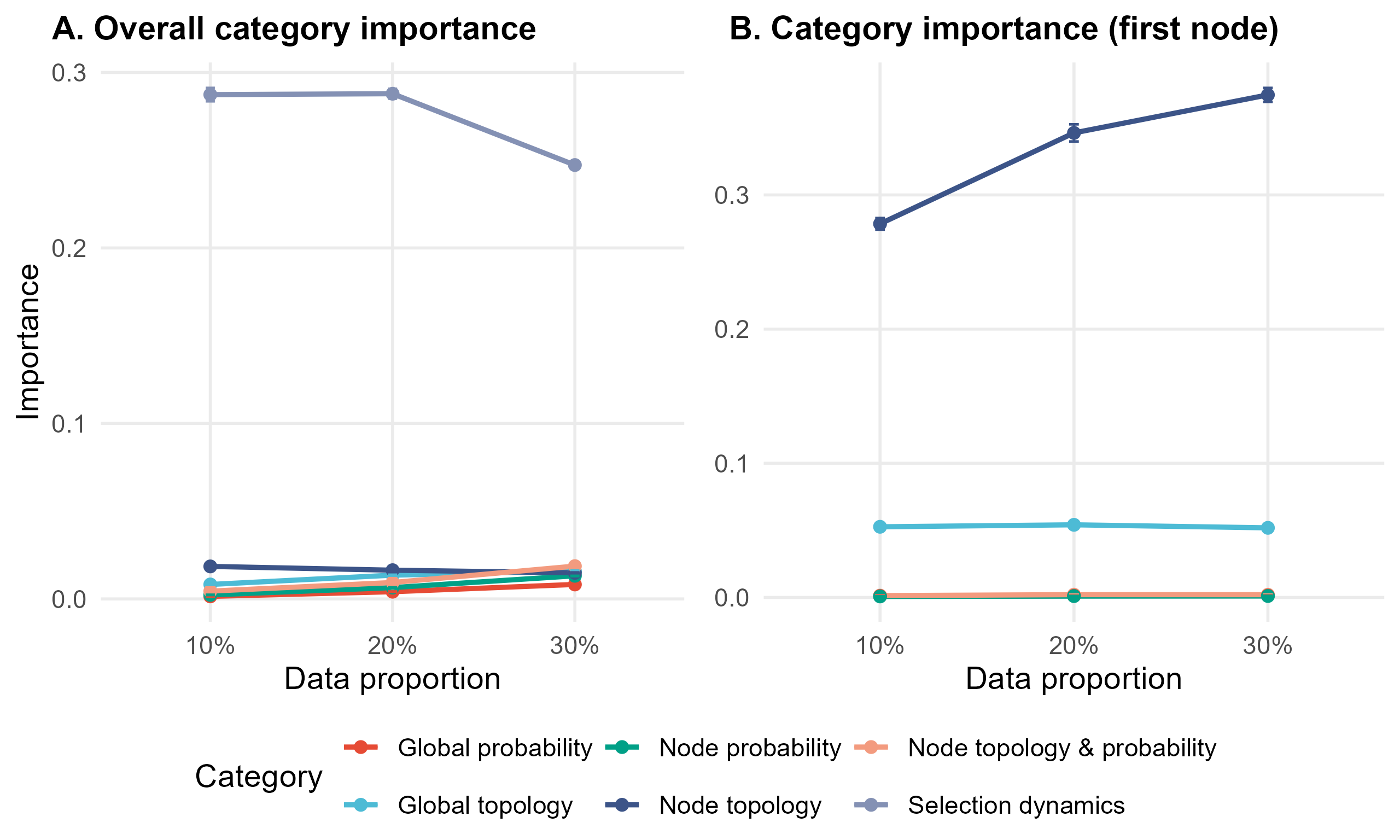

**Figure S18. Sensitivity of category-level feature importance to data proportion.** (A) Grouped permutation importance of feature categories under different proportions of training data (10%, 20%, 30%). (B) Category-level importance for the first selected node (rank = 1) under different data proportions. Bars represent the mean permutation importance scores across 100 repetitions, with error bars showing the standard deviations.

Figure.S19

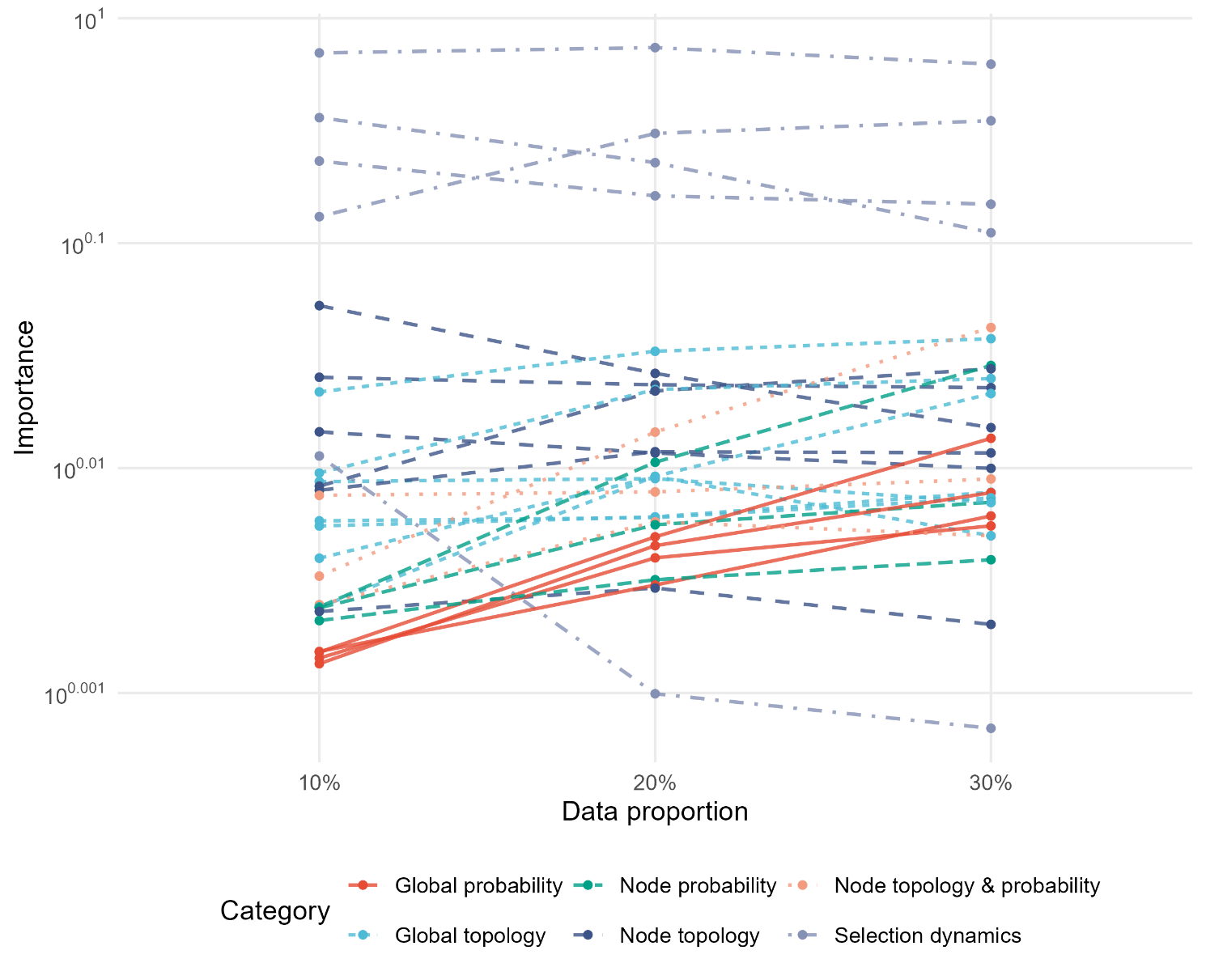

**Figure S19. Sensitivity of feature-level importance to data proportion.** Permutation importance of individual features (colored by category) under different proportions of training data (10%, 20%, 30%). Bars represent the mean permutation importance scores across 100 repetitions.
